# Association of serum antibody to serotype-specific capsular (K), lipopolysaccharide (O) and MrkA with risk reduction of invasive *Klebsiella pneumoniae* disease in young infants: an observational study

**DOI:** 10.64898/2026.07.10.26357734

**Authors:** Alane Izu, Ziyaad Dangor, Anne Amulele, Moreka Ndumba, Agnes Ndirangu, Vicky Baillie, Caroline Tigoi, James A. Berkley, Martina Carducci, Luca Rovetini, Gianina Florentina Belciug, Nicole Benson, Nicholas Dean, Francesca Micoli, Usman N. Nakakana, Courtney Olwagen, Heena Ranchod, Omar Rossi, Shabir A. Madhi

**Author notes:** **Corresponding author:** Prof Shabir Madhi Wits-VIDA, 11th Floor, Central-West Wing, Chris Hani Baragwanath Academic Hospital Chris Hani Road, Soweto, 2013 South Africa. In a case–control study, African infants under 90 days age with invasive *Klebsiella pneumoniae* disease had lower serum serotype-specific polysaccharide (K) IgG to the homotypic K-antigen, and lower MrkA IgG, compared with controls, suggesting these as potential vaccine antigens. Funding sources* The original samples from Kenya were collected under core grant support to KWTRP from Wellcome Trust (203077/Z/16/Z) and additional work was funded by Gates Foundation (INV041685 to JAB). Sample collection from South Africa was funded by Wits-VIDA. All immune marker testing performed by GVGH funded by Gates Foundation (pp). Disclosure statement* ZD and SM report grants from Gates Foundation to WITS-VIDA. AA and JAB reports funding from Gates Foundation and WELLCOME trust to their institution to support this work. FM reports partial funding from Gates Foundation for experiments present in this work. UNN and NB report grants from Gates Foundation to WITS-VIDA for this work and are employees of Gates Foundation. JAB reports grants from EDCTP and GARDP to their institution. CO reports grants from Gates Foundation and consulting fees from WHO as well as support to attend meetings from both Gates Foundation and WHO. SM reports grants Pfizer and Minervax to WITS-VIDA and clinical trial funding to institution from MERK. SM reports honoraria for lecture from Sanofi and Pfizer. SM serves on DSMB on mPox vaccine in pregnant women for Bavarian Nordic. FM and OR hold shares of GSK; FM, MC and OR hold share options of GSK; and FM, GB, LR, MC and OR are GSK employees.

## Abstract

**Background:** *Klebsiella pneumoniae* is a leading cause of sepsis in young infants. We evaluated the association of invasive *K. pneumoniae* disease (iKPnD) in relation to antigen-specific immunoglobulin G (IgG) and serum bactericidal activity (SBA) to four polysaccharide capsular (K) serotypes and five lipopolysaccharide (O) serotypes, as well as IgG to MrkA, in infants less than 90 days of age.

**Methods:** We conducted a retrospective case-control study in Kenyan and South African infants with blood culture-confirmed iKPnD. Serotype-specific antigen IgG concentrations of cases were compared with hospitalised controls without iKPnD. Geometric mean concentrations (GMCs) were estimated, and scaled covariate-adjusted models were used to estimate risk reduction over a grid of antibody concentrations.

**Results:** Transplacental transfer of IgG against various *K. pneumoniae* antigens increased with advancing gestational age. Serum IgG GMCs (expressed in RLU/mL) to disease-causing homotypic K- or O-serotypes were lower in cases compared with controls for anti-K2 (396 [95%CI: 250-628] vs 660 [95%CI: 562-776]), anti-K25 (396 [95%CI: 251-623] vs 1170 [95%CI: 988-1385]), anti-K149 (327 [95%CI: 204-521] vs 492 [95%CI: 435-557]); as well as anti-O1αβ,2α IgG (1282 [95%CI: 782-2101] vs 2250 [95%CI:1904-2658]). Furthermore, overall anti-MrkA IgG was lower in cases (945; 95%CI: 757-1179) compared with controls (1610; 95%CI: 1378-1880). SBA titres (expressed as IC50) did not differ between case and controls by K types, but were lower for O1αβ,2β in cases (27; 95%CI: 12-63 vs. 136; 95% CI: 84-221).

**Conclusion:** Our findings provide preliminary evidence that low antibodies against three of four K-antigens, O1αβ,2β and MrkA are inversely associated with iKPnD, and should be explored as potential vaccine antigens.

## Introduction

Globally, *Klebsiella pneumoniae* is a leading cause of mortality among children under-5 years of age; including being attributed in the causal chain to death in 21% of all decedents under-5 years of age in low- and middle-income countries.^1^ A vaccine administered to pregnant women could confer protection against iKPnD disease in the early-infancy period via transplacental antibody transfer.^2^

Potential *K. pneumoniae* vaccine antigen targets include polysaccharide capsular (K), lipopolysaccharide (O), and conserved protein antigens.^3^ The polysaccharide capsule of *K. pneumoniae* is a key virulence factor, enabling host phagocytic cell evasion. Although the K-antigen is highly immunogenic, it is genetically diverse with >180 capsular loci identified,^4^ with high geographic and temporal variability in the K-serotypes causing iKPnD.^5–9^ Approximately 50% of iKPnD in neonates are caused by five K-types. In contrast, the O-antigen is more restricted in genetic diversity (n=14 serotypes),^5^with a quadrivalent O-antigen vaccine potentially covering more than 85% of strains causing iKPnD.^5^ Furthermore, there is extensive IgG cross-reactivity among O-antigens with similar chemical composition.^10^ The MrkA protein, the major subunit of type 3 fimbriae, is a highly conserved adhesin critical for biofilm/abiotic surface adherence and prevalent in about 88% of invasive disease isolates, and is also a potential vaccine antigen.^11–14^ To date, there are no licensed vaccines against *K. pneumoniae*.

Vaccine antigen selection can be informed through sero-epidemiology studies evaluating for serological thresholds of risk reduction (SToRR) for disease, as done for *Neisseria meningitidis, Haemophilus influenzae* type b, and *Streptococcus agalactiae* (Group B streptococcus; GBS).^15–19^

We evaluated the association between antigen-specific immunoglobulin G (IgG) to four K-serotypes (K2, K25, K102, K149), five O-serotypes (O1αβ,2α, O1αβ,2β, O2α, O2β, and O5) and MrkA in relation to risk of iKPnD in young infants. Also, we evaluated the association of risk of iKPnD in relation to serum bactericidal activity (SBA) directed against the four K-serotypes. Furthermore, we investigated the association of gestational age (GA) on the transplacental transfer of serotype-specific IgG from mother to newborn.

## Methods

### Study design

We undertook a retrospective, observational, case-control study that included infants less than 90 days of age with culture-confirmed iKPnD and compared their serotype-specific IgG levels against the homotypic serotype of the infecting strain to that of hospitalised counterparts who remained free of iKPnD. The cases and controls had been previously enrolled in large observational studies undertaken in South Africa^19–21^ and Kenya.^21,22^

In South Africa, the decision to investigate for invasive bacterial disease, including blood culture or cerebrospinal fluid (CSF) culture, was made at the discretion of the attending physician. The dominant K-loci identified in the 226 invasive isolates sequenced in South Africa were KL25 (n=49; 21.7%), KL2 (n=40; 17.7%), KL149 (n=33; 14.6%), and KL102 (n=28; 12.4%) (unpublished data).

In Kenya, surveillance for bacteraemia has been conducted in children admitted to a hospital in Kilifi since 1998.^23^ Blood cultures are conducted on all admissions except those admitted for elective surgery or minor injuries. Blood or CSF cultures were performed at clinicians’ discretion. Of the 270 invasive isolates identified in age-eligible infants between January 2001 and April 2023, the dominant K-loci in Kenya identified by whole genome sequencing were KL2 (n=28, 9%), KL25 (n=27, 8.7%) and KL102 (n=25, 8.1%).

Further details on study design, surveillance and sample storage are described in the supplementary material.

Preferably, controls should be individuals with similar exposure to the specific disease-causing serotype; however, the available controls in our analyses had not been evaluated for *K. pneumoniae* colonization. Nevertheless, epidemiological studies from South Africa and Kilifi in Kenya report that approximately 70% of newborns hospitalised for more than 72 hours have gastrointestinal colonization by *K. pneumoniae*.^24^ Consequently, a random sample of controls who were matched to cases (4:1 where possible) were selected from South Africa and Kenya. The controls were matched to iKPnD cases according to birth weight bands (<1500 grams, 1500-2499 grams, and >2500 grams), length of hospital stay (<72 hours, 4 to <15 days, and >= 15 days), and date of birth within 60 days of the corresponding case. The matching on date of birth increases the likelihood that controls had similar strain-specific exposure compared with cases within the same facility.

For this study, only iKPnD cases of KL2, KL25, KL102 and KL149 strains were selected, for which the corresponding O-serotypes were O1αβ,2α, O1αβ,2β, O2β, O4, and O5.^25^

All serum samples were shipped on dry ice to GSK Vaccines Institute for Global Health (GVGH) in Siena, Italy, for serological testing.

### Laboratory methods

Serological measurements evaluated serum serotype-specific IgG to K2, K25, K102, K149, O1αβ,2α, O1αβ,2β, O2α, O2β, and O5 antigens and MrkA.^26^ Serum bactericidal activity (SBA) was assessed against clinical isolates harbouring KL2 (O1αβ,2β), KL25 (O5), KL102 (O2β), and KL149 (O4) which were sourced from the Wits-VIDA invasive isolates. Antigen specific IgG were assessed against each of the ten antigens for all samples, whereas SBA titres were obtained by testing cases sera and the corresponding matched controls only against the strain matched for the cases’ KL type. Tested samples for both cases and controls came from cord blood if available, otherwise serum from infant at time of event were used. Additionally, all Luminex and SBA assays were performed on maternal samples where available.

The methods for the K- and O- serotype specific IgG and MrkA IgG, as well as SBA has been previously described and a detailed description is provided in the supplementary material.^10,26^ Both the multiplex bead-based assay and the various SBA assays were characterised before the analysis in terms of accuracy, linearity, specificity, precision, limit of detection and qualification. The units of measurement and lower limit of quantification (LLoQ) for measuring IgG and SBA is detailed in **Error! Reference source not found.**.

Results of the SBA were expressed as reciprocal serum dilution that resulted in a 50% reduction of luminescence (IC50), corresponding to 50% growth inhibition of the bacteria present in the assay. GraphPad Prism software (GraphPad Software, La Jolla, CA, USA) was used for curve fitting and IC50 determination.

### Statistical methods

A statistical analysis plan was completed prior to the analysis, which is summarised in **Error! Reference source not found..**

To assess the association between birth weight and IgG as well as cord-maternal ratio of IgG (CMR), a simple linear regression model was fit to the log values of IgG. Association was determined if there was a significant change in IgG associated with a change in explanatory variable (p-value <0.05). The same analysis was done to assess the association between GA and IgG (and CMR); which was restricted to South Africa where all cases and 180 controls had documented GA. The analysis of efficiency of transplacental IgG transfer was limited to mother-newborn dyads with available paired cord-maternal blood samples and there were detectable IgG in both samples.

To assess the association between disease and immunological measurements (IgG and SBA), the focus was on cord and infant samples to evaluate protection against iKPnD in the infant. Empirical distribution of infant IgG for case and control was plotted, and receiver operating characteristic (ROC) curves were derived for infant IgG. Geometric mean concentrations (GMCs) and geometric mean titres (GMTs) of infant IgG (along with 95% confidence intervals), as well as SBA in cases and controls, were analysed.

Only one maternal–infant dyad had IgG values below the lower limit of quantification (LLoQ) (K2 only), with both maternal and infant samples below the LLoQ. A higher proportion of samples had values below the LLoQ for SBA. Analyses included values below the LLoQ (assigned half the LLoQ). For SBA analyses were done including as well as excluding values below the LLoQ.

The relative risk reduction (RRR_a_) for a given assay value, is defined as the probability of a clinical endpoint occurring at assay value, divided by the risk of a clinical endpoint occurring at the lowest assay value (typically the lower limit of detection). A covariate-adjusted logistic regression model (CALM),an extension of the scaled logit model, was used to estimate the RRR_a_ for possible assay values.^27,28^ Further details of the model are described in the supplementary material.

Correlation between IgG and SBA is reported including all available samples, and also when excluding samples where IgG or SBA were below the LLoQ.

All analyses were done including only cases with the homotypic serotype (e.g. when assessing anti-K2 IgG, only cases with strains with K locus (KL) 2 were used in the analyses). Considering potential antibody cross-reactivity with O1αβ,2α and O1αβ,2β, we also compared pooled cases with either O1αβ,2α and O1αβ,2β strains to controls.^25,29^

To investigate the impact of timing of sample collection on the association iKPnD and immune response, a sensitivity analysis to investigate the association between IgG and iKPnD limited to only cord blood samples was performed.

### Ethics approval

This study was approved by the University of the Witwatersrand Human Ethics Research Committee and by KEMRI Scientific and Ethic Review (SERU). Details on ethics approval and participant consent is provided in supplementary material.

## Results

A total of 98 infant iKPnD cases and 292 matched controls were analysed; Table 1. The majority of iKPnD cases (74.5%; 73/98) and controls (85.6%,250/292) were from South Africa. Except for five South African cases in which *K. pneumoniae* was identified from CSF, the rest of the cases were cultured from blood. The KL distribution of cases included in the study were: KL25 (n=33), KL2 (n=28), KL102 (n=22), and KL149 (n=15). All the KL149 cases were from South Africa. The distribution of O-serotypes amongst cases were O5 (n = 32), O1αβ,2β (n = 25), O2β (n = 22), O1αβ,2α (n = 16), and O4 (n = 3). The O-types associated with KL2 were O1αβ,2α (15/28), O1αβ,2β (12/28) and O2β (1/28). In contrast, KL25 was nearly uniformly associated with O5 (32/33), KL102 with O2β (21/22), and KL149 with O1αβ,2β (12/15). The three O4 invasive isolates were all associated with KL149; Table1.

**Table 1:** Number of available and tested samples from infants who developed invasive *Klebsiella pneumoniae* disease (iKpnD).

| Classification | KE | SA (CHBAH) | SA (RMMCH) | Total |
| --- | --- | --- | --- | --- |
| **KL-type** |  |  |  |  |
| KL2 | 11 (8/3)* | 15 (10/5) | 2 (1/1) | 28 (19/9) |
| O1αβ,2α | 9 (7/2) | 5 (3/2) | 1 (1/0) | 15 (10/5) |
| O1αβ,2β | 2 (1/1) | 9 (6/3) | 1 (0/1) | 12 (8/4) |
| O2β | 0 (0/0) | 1 (1/0) | 0 (0/0) | 1 (1/0) |
| KL25 | 9 (8/1) | 20 (12/8) | 4 (2/2) | 33 (22/11) |
| O1αβ,2α | 0 (0/0) | 1 (1/0) | 0 (0/0) | 1 (1/0) |
| O5 | 9 (8/1) | 19 (11/8) | 4 (2/2) | 32 (21/11) |
| KL102 | 5 (5/0) | 17 (15/2) | 0 (0/0) | 22 (20/2) |
| O1αβ,2β | 0 (0/0) | 1 (1/0) | 0 (0/0) | 1 (1/0) |
| O2β | 5 (5/0) | 16 (14/2) | 0 (0/0) | 21 (19/2) |
| KL149 | 0 (0/0) | 14 (12/2) | 1 (1/0) | 15 (13/2) |
| O1αβ,2β | 0 (0/0) | 11 (10/1) | 1 (1/0) | 12 (11/1) |
| O4 | 0 (0/0) | 3 (2/1) | 0 (0/0) | 3 (2/1) |
| **O-type** |  |  |  |  |
| O1αβ,2α | 9 (7/2) | 6 (4/2) | 1 (1/0) | 16 (12/2) |
| O1αβ,2β | 2 (1/1) | 21 (17/4) | 2 (2/0) | 25 (20/5) |
| O2β | 5 (5/0) | 17 (15/2) | 0 (0/0) | 22 (20/2) |
| O4 | 0 (0/0) | 3 (2/1) | 0 (0/0) | 3 (2/1) |
| O5 | 9 (8/1) | 19 (11/8) | 4 (2/2) | 32 (21/11) |
| **Controls** | 42 (30/12) | 229 (229/0) | 21 (21/0) | 292 (280/12) |
| **SBA K2 (O1αβ,2β)** |  |  |  |  |
| Cases | 11 (8/3) | 15 (10/5) | 2 (1/1) | 28 (19/9) |
| Controls | 18 (16/2) | 60 (60/0) | 8 (8/0) | 86 (84/2) |
| **SBA K25 (O5)** |  |  |  |  |
| Cases | 10 (8/2) | 20 (12/8) | 4 (2/2) | 34 (22/12) |
| Controls | 12 (5/7) | 68 (68/0) | 9 (9/0) | 89 (82/7) |
| **SBA K102 (O2β)** |  |  |  |  |
| Cases | 5 (5/0) | 17 (15/2) | 0 (0/0) | 22 (20/2) |
| Controls | 13 (9/4) | 63 (63/0) | 0 (0/0) | 76 (72/4) |
| **SBA K149 (O4)** |  |  |  |  |
| Cases | 0 (0/0) | 14 (12/2) | 1 (1/0) | 15 (13/2) |
| Controls | 0 (0/0) | 38 (38/0) | 4 (4/0) | 42 (42/0) |

\*Total samples are shown and the breakdown of cord or acute samples are shown in parenthesis (cord/acute). IgG binding assays were performed on all cases and control samples. Serum Bactericidal Assay (SBA) was performed on a subset of cases and controls. For cases, SBA for the homologous K-type was done and for controls, SBA for the K-type of the matched case was performed.

The median length of hospital stay for cases and controls was 7 days (IQR: 3-16 days); Table 2. Among iKPnD cases, 51% (50/98) were born at birth weight of <1500 grams and 35% (34/98) were from 1500 and 2499 grams. Among cases with known GA, 70.0% (56/80) were born at <34 weeks, including 55% (44/80) at <32 weeks; 16.2% (13/80) were born at term. GA was unknown for 18.4% (19/98) of cases, all from Kenya. Approximately, one-third of cases demised (35/98) compared with 15% (43/292) of controls.

**Table 2:** Demographic characteristics of participants.

| Variable | Overall | Case<sup>1</sup> | Control |
| --- | --- | --- | --- |
| Total | 390 | 98 | 292 |
| Kenya | 67 (17.2) | 25 (25.5) | 42 (14.4) |
| South Africa | 323 (82.8) | 73 (74.5) | 250 (85.6) |
| Median Length of stay -days (IQR) | 7 (3-16) | 7 (4-14.75) | 7.5 (3-16) |
| 0-<3 | 58 (14.9) | 18 (18.4) | 40 (13.7) |
| 3-<15 | 211 (54.1) | 55 (56.1) | 156 (53.4) |
| >=15 | 112 (28.7) | 25 (25.5) | 87 (29.8) |
| Unknown length of stay | 9 (2.3) | 0 (0.0) | 9 (3.1) |
| Median Birth weight -grams (IQR) | 1605 (1190-2230) | 1445 (1115-1955) | 1700 (1210-2326.5) |
| <1500 | 164 (42.1) | 50 (51.0) | 114 (39.0) |
| 1500-<2500 | 155 (39.7) | 34 (34.7) | 121 (41.4) |
| >=2500 | 70 (17.9) | 14 (14.3) | 56 (19.2) |
| Unknown birthweight | 1 (0.3) | 0 (0.0) | 1 (0.3) |
| Median Gestational age -weeks (IQR) | 32 (29-36) | 31 (28-35) | 33 (30-37) |
| <32 | 117 (44.7) | 44 (55.0) | 73 (40.1) |
| 32-<34 | 34 (13.0) | 12 (15.0) | 22 (12.1) |
| 34-<37 | 49 (18.7) | 11 (13.8) | 38 (20.9) |
| >=37 | 62 (23.7) | 13 (16.2) | 49 (26.9) |
| Unknown gestational age | 128 (32.8) | 18 (18.4) | 110 (37.7) |
| Outcome |  |  |  |
| Discharged | 299 (77.5) | 63 (64.3) | 236 (81.9) |
| Transferred | 9 (2.3) | 0 (0.0) | 9 (3.1) |
| Died | 78 (20.2) | 35 (35.7) | 43 (14.9) |
| Unknown outcome | 4 (1.0) | 0 (0.0) | 4 (1.4) |

<sup>1</sup>All cases were identified from bloodstream infections, except for 5 South African cases and 1 Kenyan case where *Klebsiella pneumoniae* was identified from CSF.

### Maternal IgG and cord-maternal ratio of IgG with birth weight and GA

Maternal serum IgG GMCs were similar among women irrespective of birth weight and GA. The CMRs increased with increasing birth weight or GA; Supplementary Figure 1a and Figure 1a. In mother-infant dyads, the log CMR for serotype-specific anti-K, anti-O, and MrkA IgG increased between 0.16 to 0.30 per 1kg increase in birth weight; Supplementary Table 4. Log CMRs in South African dyads were 0.10 to 0.32 lower than in Kenyan counterparts across all K- and O- serotype-specific IgG, as well as MrkA IgG; Supplementary Table 4. For newborns weighing >2500g, the expected CMR in Kenya ranged from 0.78 to 1.12, compared with 0.68 to 0.87 in South Africa, albeit 95%CI overlapped; Supplementary **Error! Reference source not found.**b. In South Africa, the log CMR for all antigen targets increased progressively from 20 weeks GA (range: 0.30-0.42 for K-antigens, 0.35-0.55 for O-antigens, and 0.38 for MrkA) to 40 weeks GA (range: 0.74-0.83 for K-antigens, 0.76-0.90 for O-antigens, and 0.85 for MrkA); Figure 1b, Supplementary Table 4.

**Figure 1:**
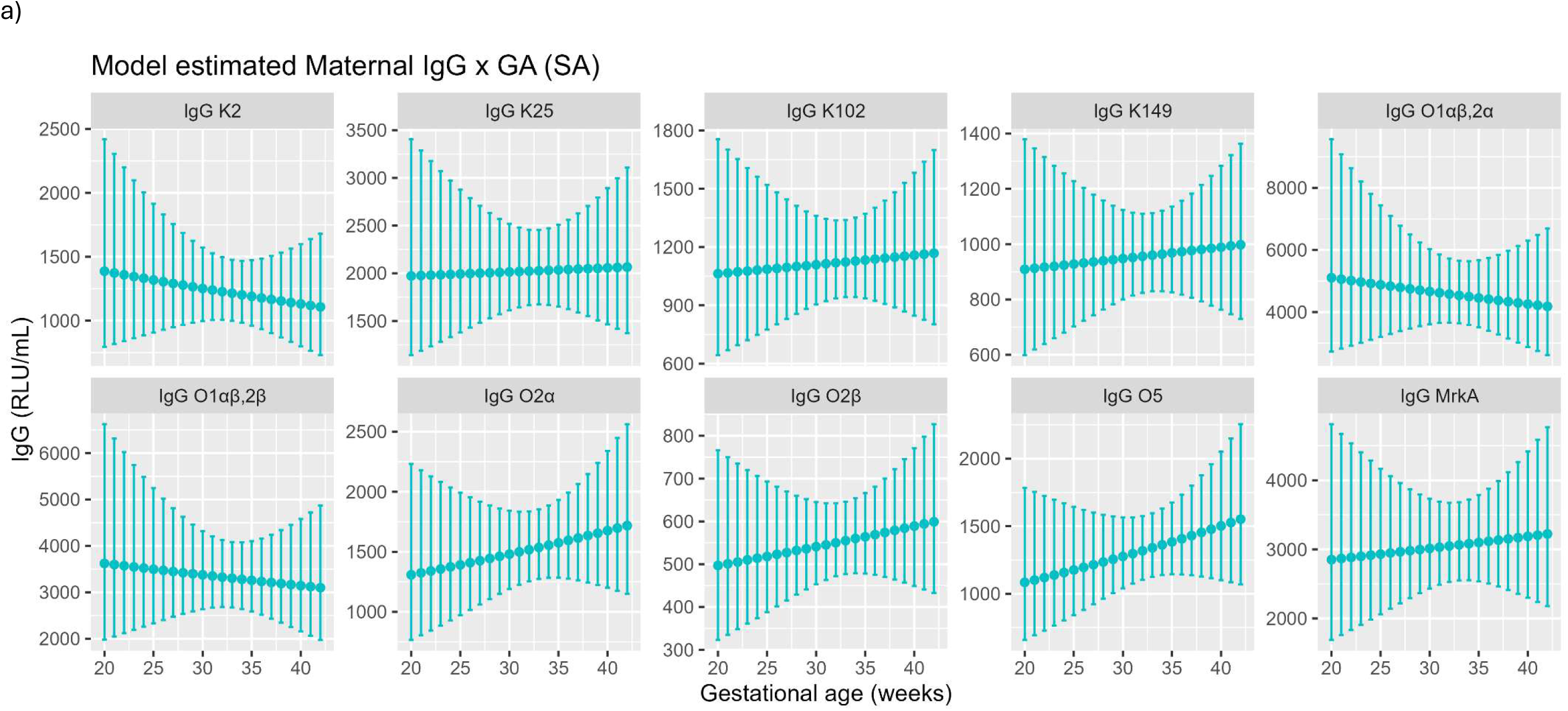
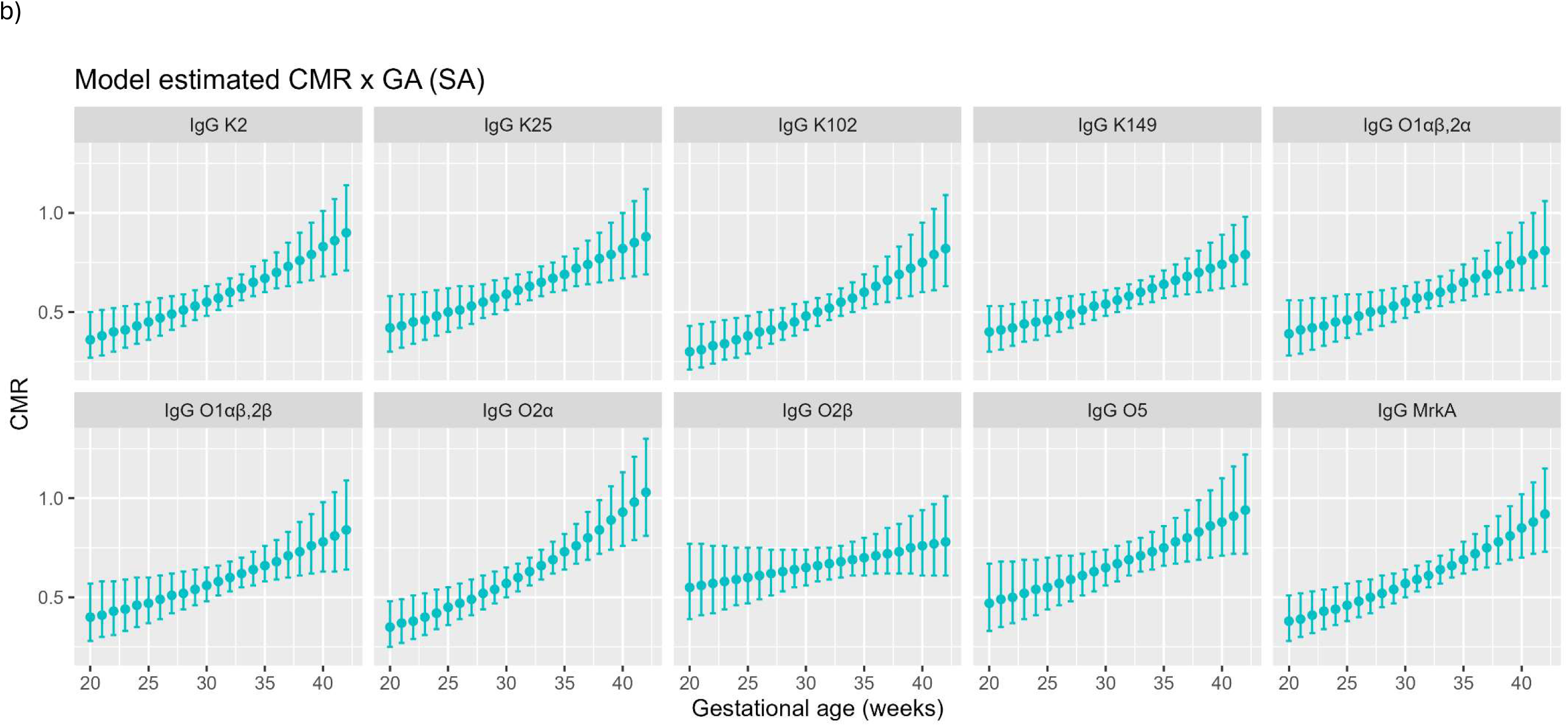
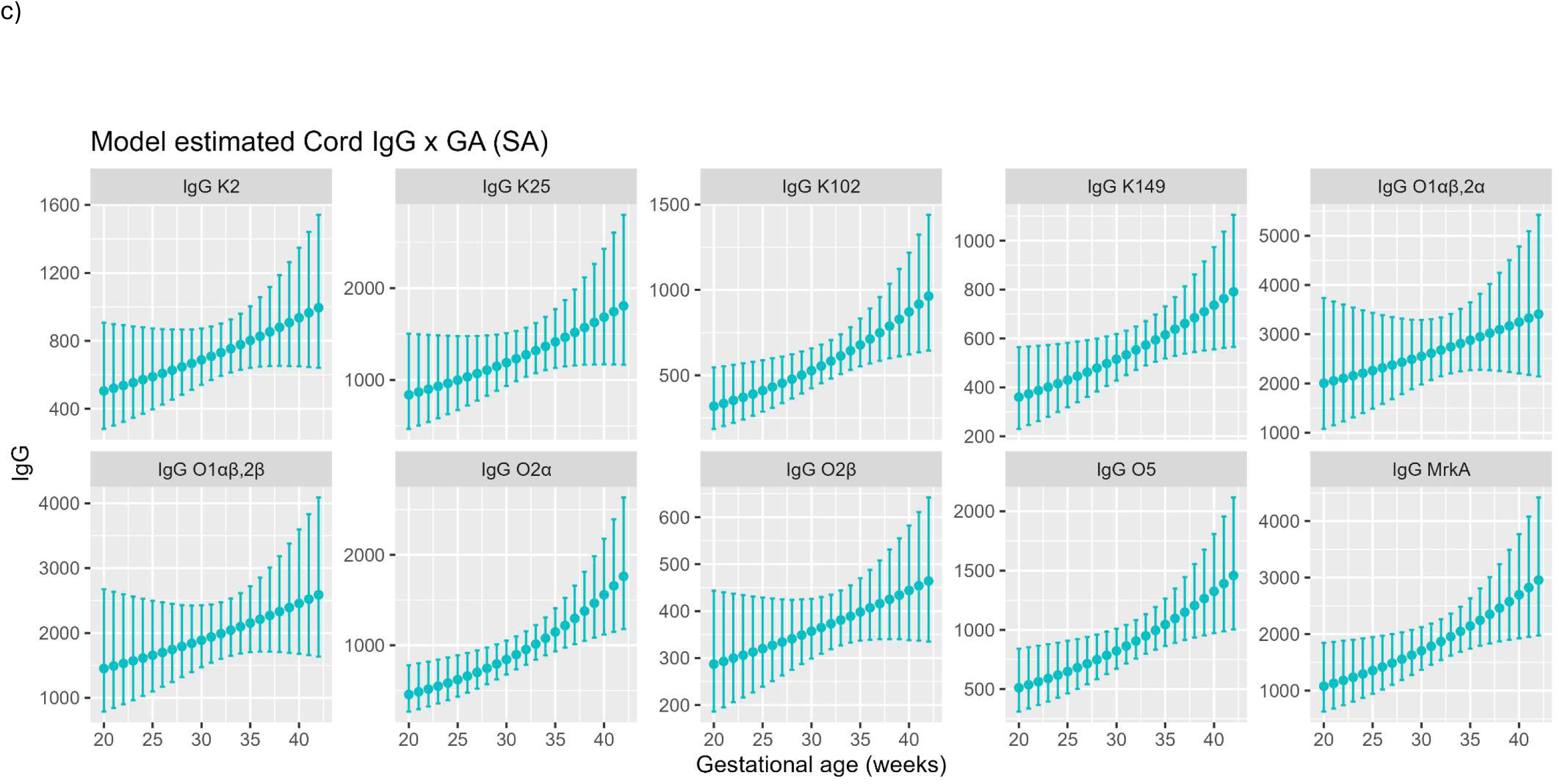
Correlation between maternal IgG, cord:maternal ratio and infant IgG with gestational age (y-axis is different for each sub-figure to highlight the trends for the different IgG). Alt text: Graphical representation of estimated maternal, Cord:maternal ratio and cord anti-K and anti-O IgG along with confidence intervals by gestational age in subfigures a to c, respectively.

### Association of infant IgG levels with birth weight and GA

Anti-K, anti-O, and MrkA IgG in cord blood from controls was modelled against birth weight and GA. Estimated parameters of the model are in Supplementary Table 3 and model estimates of IgG at various VA and birth weight are shown in Figure 1c and **Error! Reference source not found.**c. Trends mirrored that of the CMR with both increasing birth weight and GA associated with higher IgG levels. For all anti-K antigens, anti-O antigens and MrkA, cord IgG was 12.3% to 34.5% higher in infants born with a birth weight greater than 2500g compared with infants born less than1500g; **Error! Reference source not found.**c. Estimated cord serotype-specific IgG titres increased between 50% to 240% between 20 and 40 weeks GA; Figure 1c.

### Association between infant IgG and SBA and invasive Klebsiella pneumoniae disease

The serotype-specific IgG GMCs (RLU/mL) in cord/infant blood were lower in cases compared with controls for anti-K2 (396, 95%CI: 250-628 vs. 660, 95%CI: 562-776; p=0.040) and anti-K25 (396, 95%CI: 251-623 vs. 1170, 95%CI 988-1385; p<0.001); with a similar trend observed for K149 (327, 95%CI: 204-521 vs. 492, 95%CI: 435-557; p=0.091). No difference in GMCs was observed for anti-K102 IgG between cases and controls; Table 3 and **Error! Reference source not found.**a.

**Table 3:** Geometric mean concentrations (and 95% confidence intervals) between cord blood or acute serum^1^ from infants with and without invasive *Klebsiella pneumoniae* disease for various anti-K, anti-O and MrkA IgG antibodies.

| Case definition | IgG | Case [RLU/mL] | Control [RLU/mL] | p-value |
| --- | --- | --- | --- | --- |
| K2 | K2 | 396 (250-628) n=28 | 660 (561-776) n=292 | 0.040 |
| K25 | K25 | 396 (251-623) n=33 | 1170 (988-1385) n=292 | <0.001 |
| K102 | K102 | 503 (295-859) n=22 | 590 (511-681) n=292 | 0.559 |
| K149 | K149 | 327 (204-521) n=15 | 492 (435-557) n=292 | 0.091 |
| O1αβ,2α | O1αβ,2α | 1224 (436-3434) n=16 | 2250 (1904-2658) n=292 | 0.236 |
| O1αβ,2β | O1αβ,2β | 1067 (642-1774) n=25 | 1736 (1478-2038) n=292 | 0.071 |
| O1αβ,2α or O1αβ,2β<sup>2</sup> | O1αβ,2α | 1282 (782-2101) n=41 | 2250 (1904-2658) n=292 | 0.035 |
| O1αβ,2α or O1αβ,2β<sup>2</sup> | O1αβ,2β | 1059 (661-1697) n=41 | 1736 (1478-2038) n=292 | 0.051 |
| O2β | O2β | 358 (255-503) n=22 | 436 (389-488) n=292 | 0.266 |
| O5 | O5 | 695 (456-1058) n=32 | 984 (861-1125) n=292 | 0.117 |
| All cases | MrkA | 945 (757-1179) n=98 | 1610 (1378-1880) n=292 | <0.001 |
| K2 | MrkA | 910 (643-1289) n=28 | 1610 (1378-1880) n=292 | 0.004 |
| K25 | MrkA | 883 (536-1455) n=33 | 1610 (1378-1880) n=292 | 0.025 |
| K102 | MrkA | 1097 (794-1515) n=22 | 1610 (1378-1880) n=292 | 0.035 |
| K149 | MrkA | 945 (481-1856) n=15 | 1610 (1378-1880) n=292 | 0.122 |
| O1αβ,2α | MrkA | 864 (485-1541) n=16 | 1610 (1378-1880) n=292 | 0.042 |
| O1αβ,2β | MrkA | 917 (596-1411) n=25 | 1610 (1378-1880) n=292 | 0.017 |
| O2β | MrkA | 1080 (782-1491) n=22 | 1610 (1378-1880) n=292 | 0.029 |
| O4 | MrkA | 702 (51-9712) n=3 | 1610 (1378-1880) n=292 | 0.420 |
| O5 | MrkA | 949 (579-1554) n=32 | 1610 (1378-1880) n=292 | 0.045 |

<sup>1</sup>Acute serum refers to the sample collected at hospital admission. <sup>2</sup>Cases with either O1αβ,2α or O1αβ,2β O-type were compared to controls.

Except for anti-K25 IgG, confidence intervals for the reverse cumulative distribution between cases and controls overlapped for IgG values less than 1500; Supplementary Figure 3. Antibody measurements associated with 80% risk reduction in iKPnD due to strains of serotypes K2, K25, and K149 were estimated as 22,396 (95%CI: 240 to <100,000), 4,333 (95%CI: 1,763-10,651), and 1,456 (95%CI: 1,264-1,677), respectively; Figure 3 and Supplementary Table 5. Over a large grid of possible anti-K102 values, no antibody concentration was associated with a risk reduction greater than 80%; Figure 3a-3d and Supplementary Table 5.

The percentage of SBA values above the LLoQ was similar between cases (60.7% to 75.8%) and controls (51.2% to 81.6%) for all K-types, except for K149, where 83.3% (69.4-91.7) of controls were above the LLoQ compared with 46.7% (95%CI: 24.8-69.9) of cases; Table 4. When comparing SBA GMTs against each K-type responses were similar between cases and controls Table 4 and Figure 4a. Also, when excluding values below the LLoQ, SBA GMTs remained comparable between cases and controls for all K-types; Supplementary Figure 4a. There was poor to modest correlation for serotype-specific IgG and SBA in cord blood of controls for K2, K25, K102, and K149 with correlations ranging from 0.026 to 0.449; and 0.102 to 0.356 when removing samples where either IgG or SBA values were below the LLoQ; Supplementary Figures 5a and 5b.

**Table 4:**
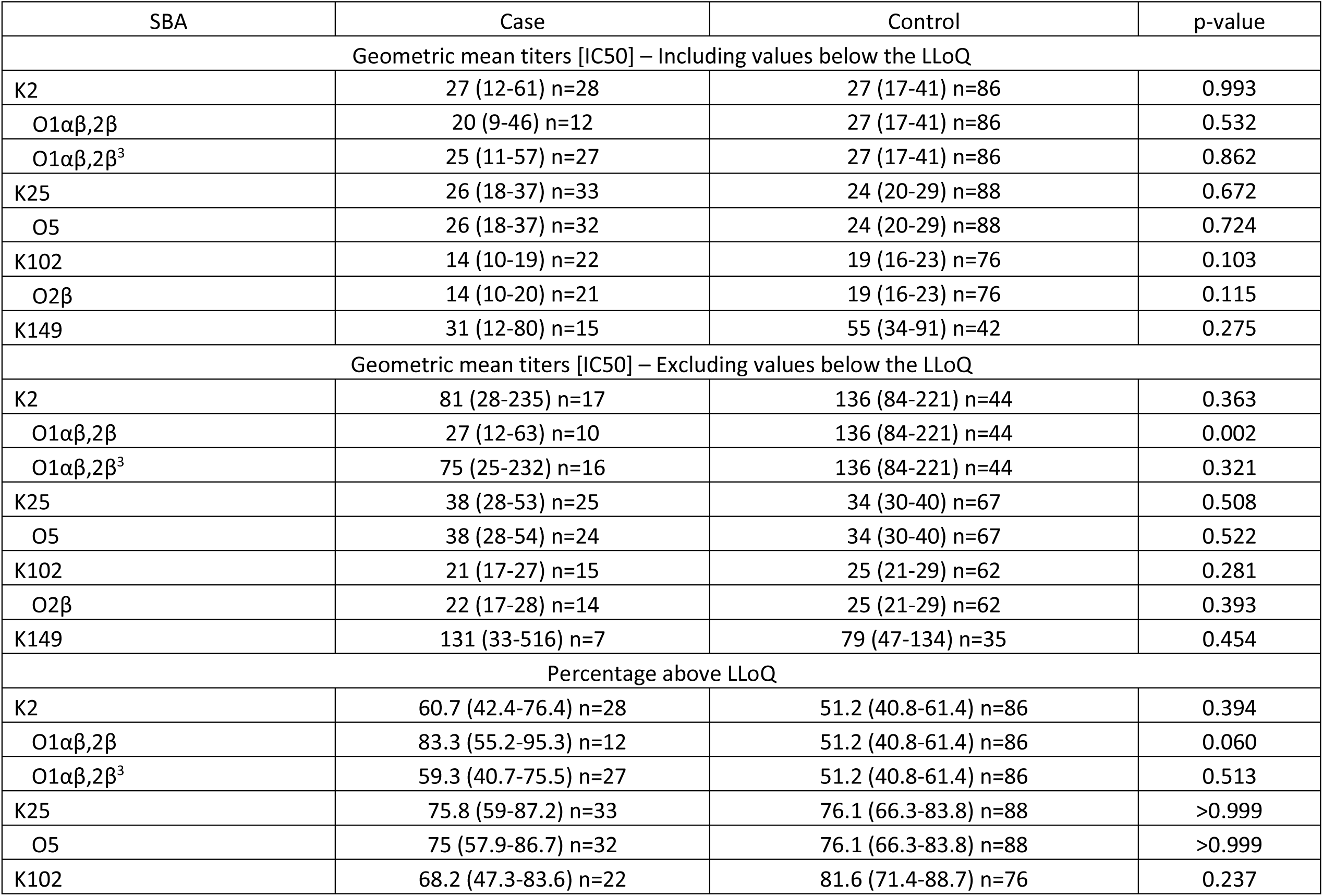
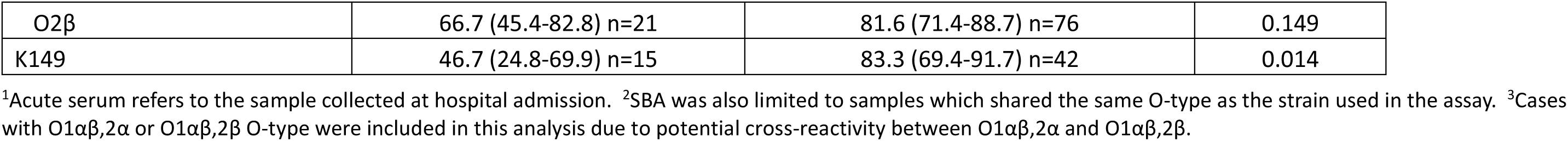
Geometric mean titers and percentage above the lower limit of quantification (LLoQ) along with 95% confidence intervals between cord blood or acute serum^1^ from infants with and without invasive *Klebsiella pneumoniae* disease for serum bactericidal activity against K2, K25, K102, K149, O1αβ,2β^2^, O2β^2^ and O5^2^. GMTs are presented when included values lower than the LLoQ (assigned half the value of LLoQ) and also when excluding values below the LLoQ were included.

| SBA | Case | Control | p-value |
| --- | --- | --- | --- |
| Geometric mean titers [IC50] – Including values below the LLoQ |  |  |  |
| K2 | 27 (12-61) n=28 | 27 (17-41) n=86 | 0.993 |
| O1$\alpha\beta$,2β | 20 (9-46) n=12 | 27 (17-41) n=86 | 0.532 |
| O1$\alpha\beta$,2β<sup>3</sup> | 25 (11-57) n=27 | 27 (17-41) n=86 | 0.862 |
| K25 | 26 (18-37) n=33 | 24 (20-29) n=88 | 0.672 |
| O5 | 26 (18-37) n=32 | 24 (20-29) n=88 | 0.724 |
| K102 | 14 (10-19) n=22 | 19 (16-23) n=76 | 0.103 |
| O2β | 14 (10-20) n=21 | 19 (16-23) n=76 | 0.115 |
| K149 | 31 (12-80) n=15 | 55 (34-91) n=42 | 0.275 |
| Geometric mean titers [IC50] – Excluding values below the LLoQ |  |  |  |
| K2 | 81 (28-235) n=17 | 136 (84-221) n=44 | 0.363 |
| O1$\alpha\beta$,2β | 27 (12-63) n=10 | 136 (84-221) n=44 | 0.002 |
| O1$\alpha\beta$,2β<sup>3</sup> | 75 (25-232) n=16 | 136 (84-221) n=44 | 0.321 |
| K25 | 38 (28-53) n=25 | 34 (30-40) n=67 | 0.508 |
| O5 | 38 (28-54) n=24 | 34 (30-40) n=67 | 0.522 |
| K102 | 21 (17-27) n=15 | 25 (21-29) n=62 | 0.281 |
| O2β | 22 (17-28) n=14 | 25 (21-29) n=62 | 0.393 |
| K149 | 131 (33-516) n=7 | 79 (47-134) n=35 | 0.454 |
| Percentage above LLoQ |  |  |  |
| K2 | 60.7 (42.4-76.4) n=28 | 51.2 (40.8-61.4) n=86 | 0.394 |
| O1$\alpha\beta$,2β | 83.3 (55.2-95.3) n=12 | 51.2 (40.8-61.4) n=86 | 0.060 |
| O1$\alpha\beta$,2β<sup>3</sup> | 59.3 (40.7-75.5) n=27 | 51.2 (40.8-61.4) n=86 | 0.513 |
| K25 | 75.8 (59-87.2) n=33 | 76.1 (66.3-83.8) n=88 | >0.999 |
| O5 | 75 (57.9-86.7) n=32 | 76.1 (66.3-83.8) n=88 | >0.999 |
| K102 | 68.2 (47.3-83.6) n=22 | 81.6 (71.4-88.7) n=76 | 0.237 |
| O2β | 66.7 (45.4-82.8) n=21 | 81.6 (71.4-88.7) n=76 | 0.149 |
| K149 | 46.7 (24.8-69.9) n=15 | 83.3 (69.4-91.7) n=42 | 0.014 |

<sup>1</sup>Acute serum refers to the sample collected at hospital admission. <sup>2</sup>SBA was also limited to samples which shared the same O-type as the strain used in the assay. <sup>3</sup>Cases with O1 $\alpha\beta$ ,2α or O1 $\alpha\beta$ ,2β O-type were included in this analysis due to potential cross-reactivity between O1 $\alpha\beta$ ,2α and O1 $\alpha\beta$ ,2β .

Serotype-specific anti-O IgG GMCs was lower in cases than controls for O1αβ,2α and O1αβ,2β, albeit not statistically significant. However, when combining O1αβ,2α and O1αβ,2β cases, anti- O1αβ,2α IgG was significantly lower in cases (1282; 95%CI: 782-2101) compared with controls (2248; 95%CI:1904-2654; p= 0.035). Also, a similar trend was observed for anti- O1αβ,2β IgG with GMCs being lower (1059; 95%CI: 661-1697) in cases (composite of O1αβ,2α and O1αβ,2β) compared with controls (1736; 95%CI: 1478-2038; p = 0.051); Table 3; Figure 2b. Although anti- O2β and -O5 IgG point estimate was also lower in cases than controls, the difference was not significant; Table 3.

**Figure 2:**
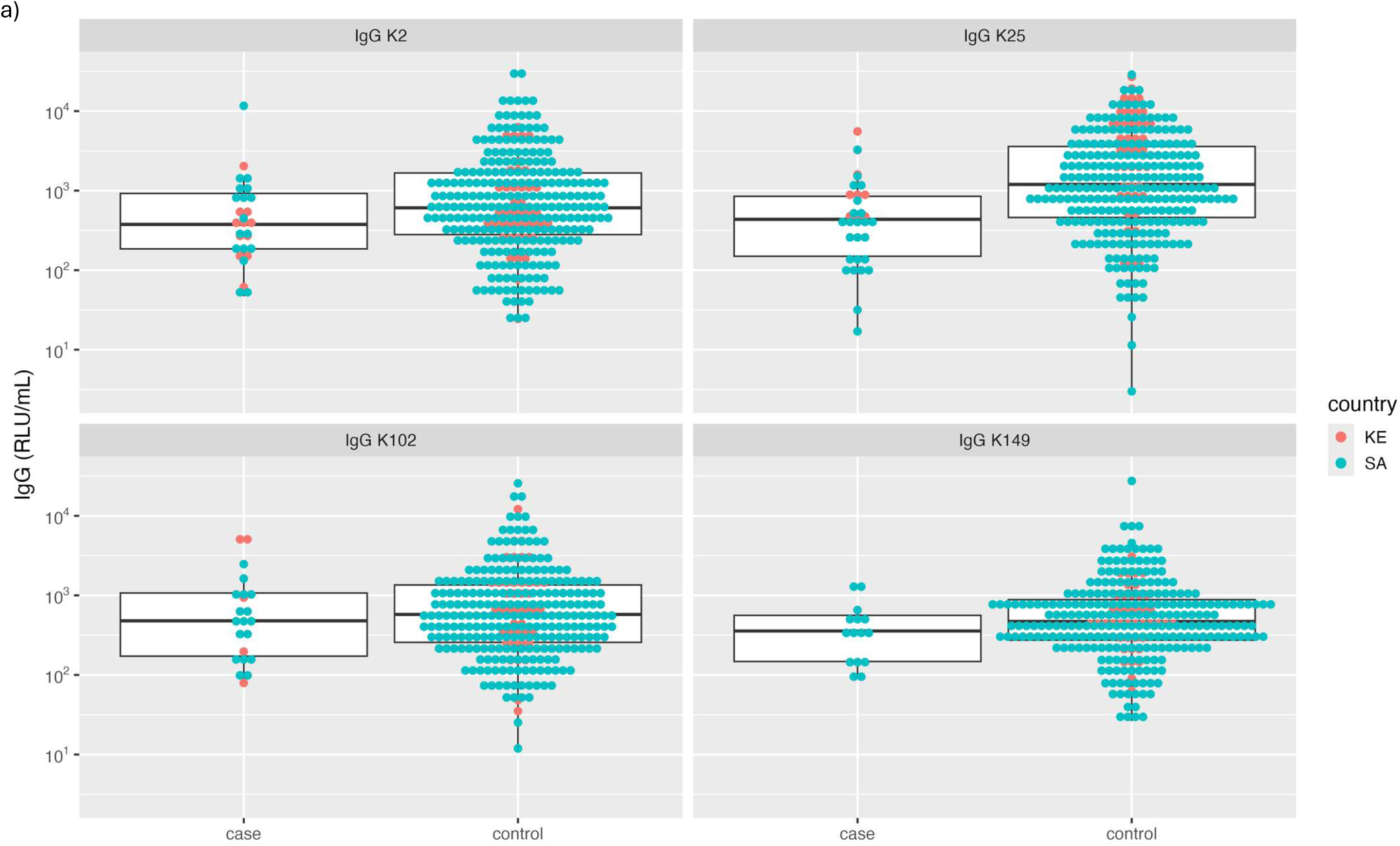
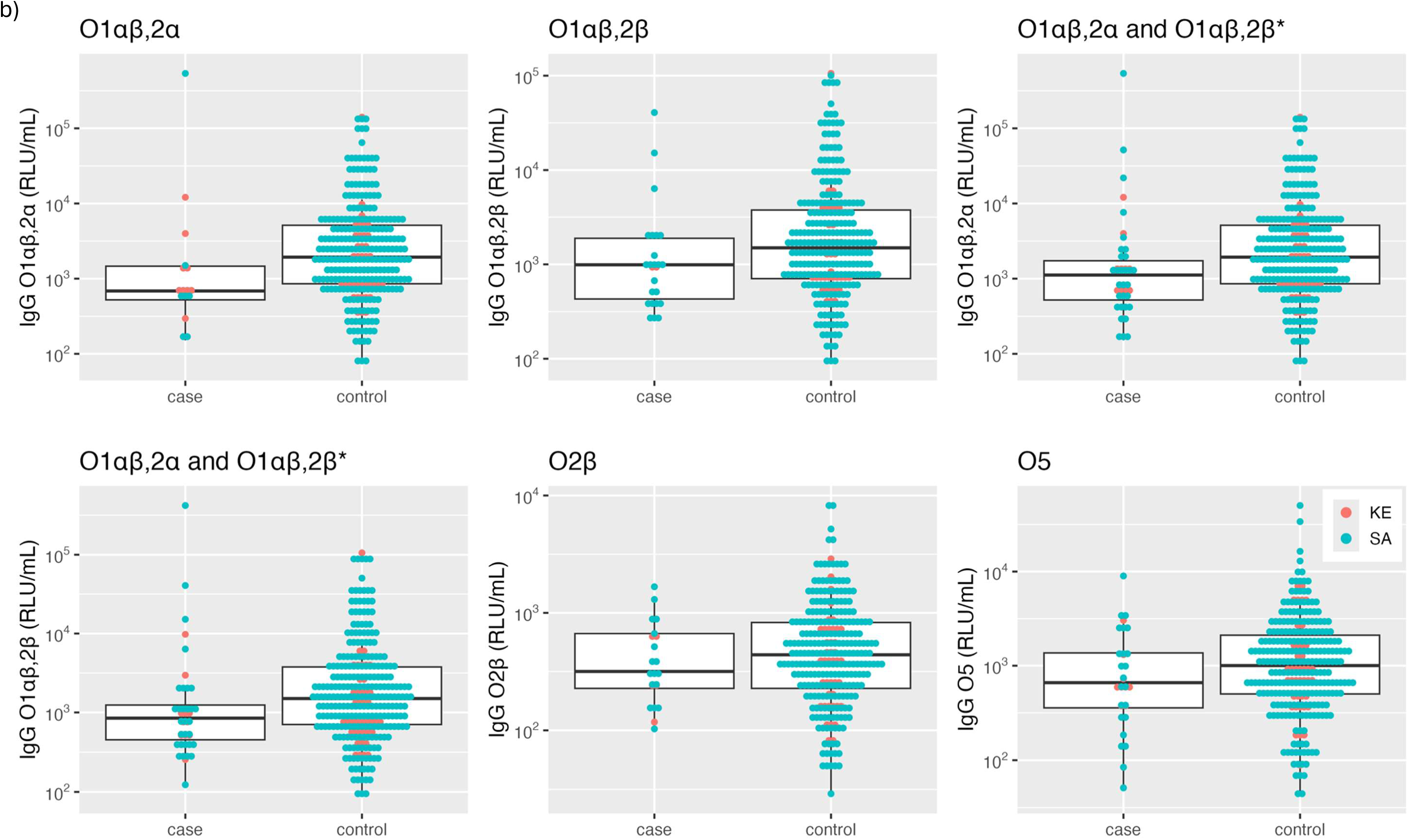
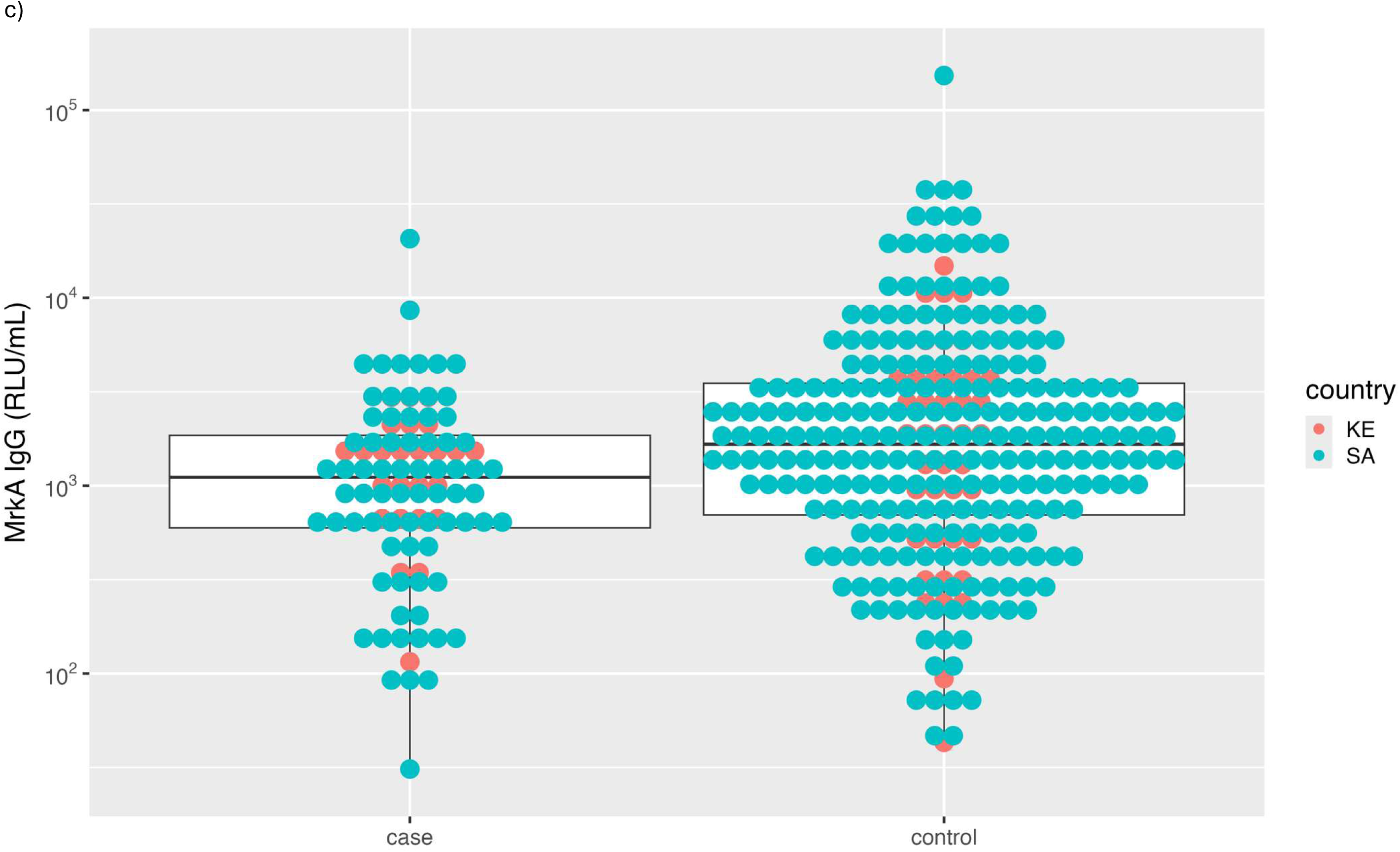
Distribution of anti-K, anti-O and MrkA IgG from infant cases and controls. Alt text: Graphical representation of anti-K, anti-O and MrkA IgG for cases and controls using dot plots with overlaying box and whisker-plots in subfigures a to c, respectively. *Figures for O1αβ,2α and O1αβ,2β display IgG for cases with O-type O1αβ,2α or O1αβ,2β and all controls.

Reverse cumulative plots are shown in Supplementary Figure 3b, where larger differences between the reverse cumulative distribution of cases and controls is seen in the combined O1αβ,2α and O1αβ,2β cases compared with controls for both anti-O1αβ,2α and anti- O1αβ,2β IgG. CALM models were fit, however, estimates of antibody measurements associated with disease risk were characterized by substantial uncertainty; Figure 3e-3h and Supplementary Table 5.

**Figure 3:**
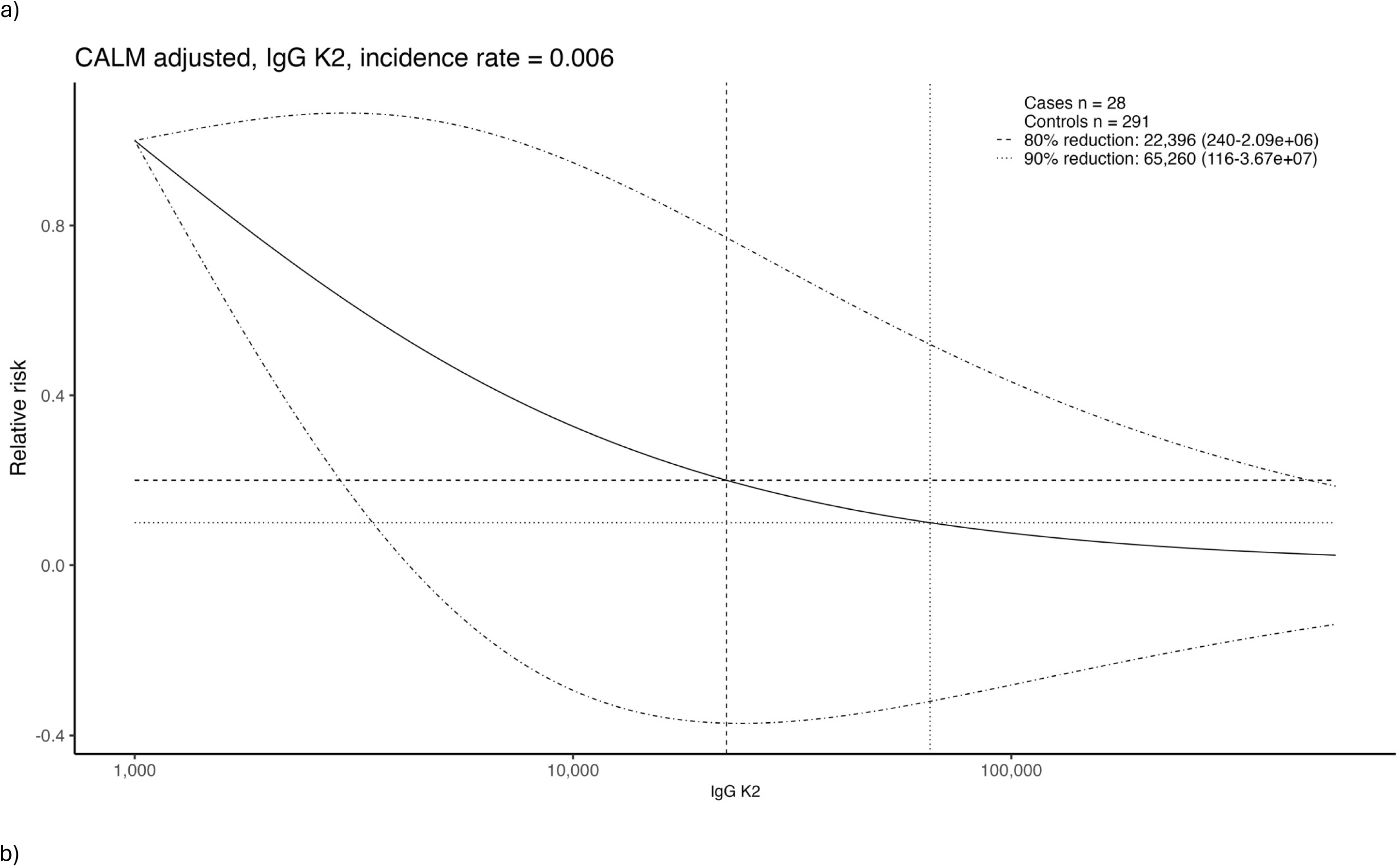
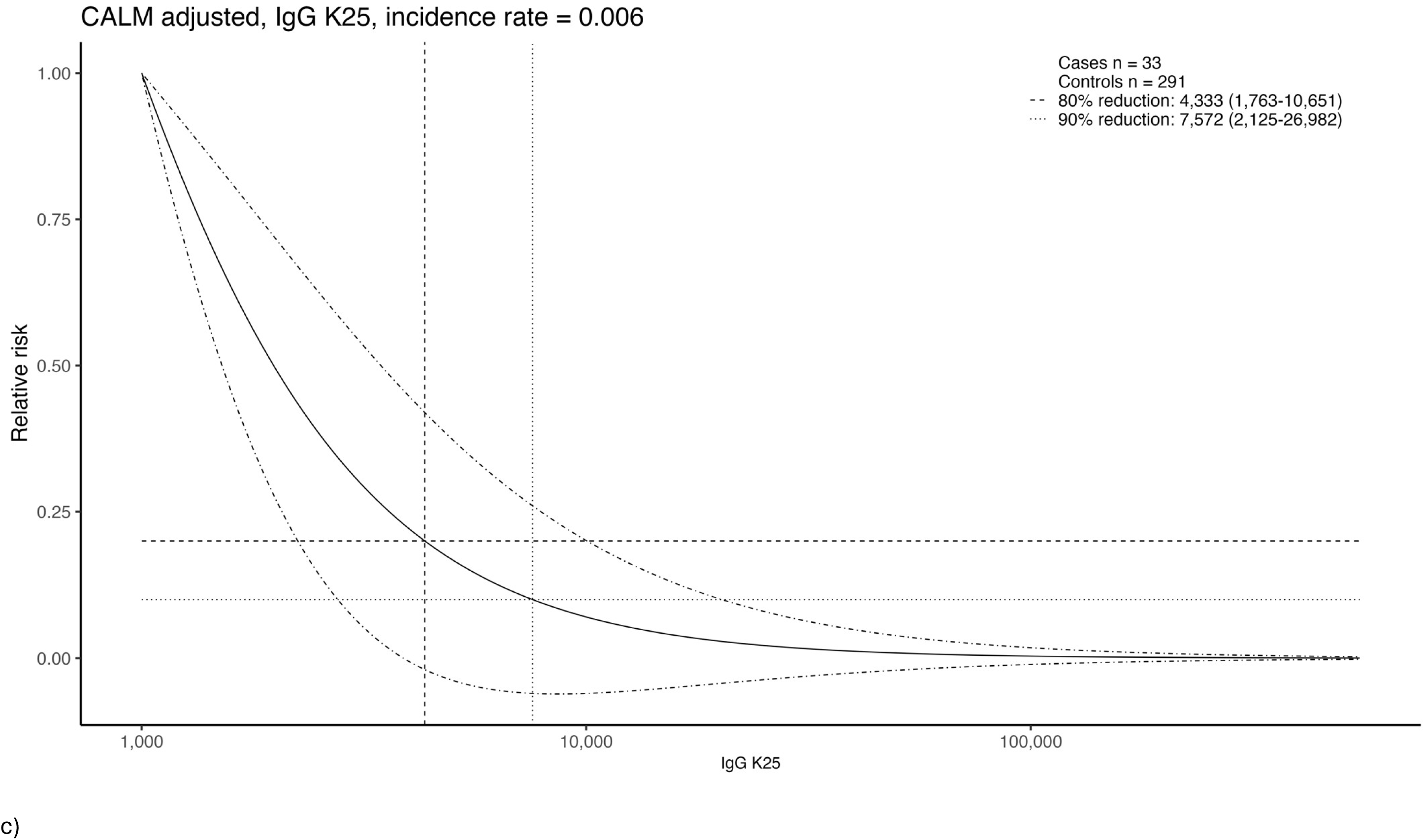
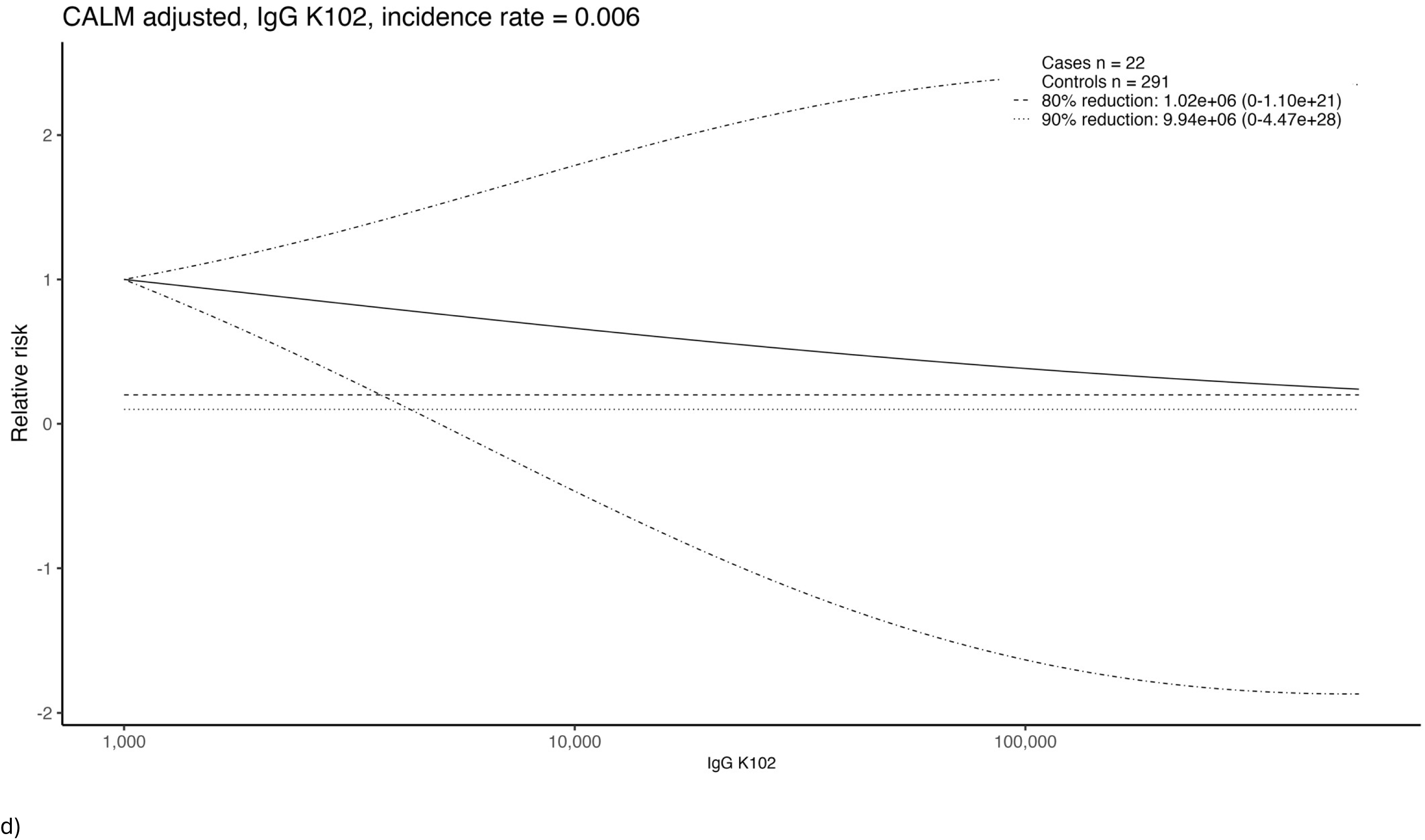
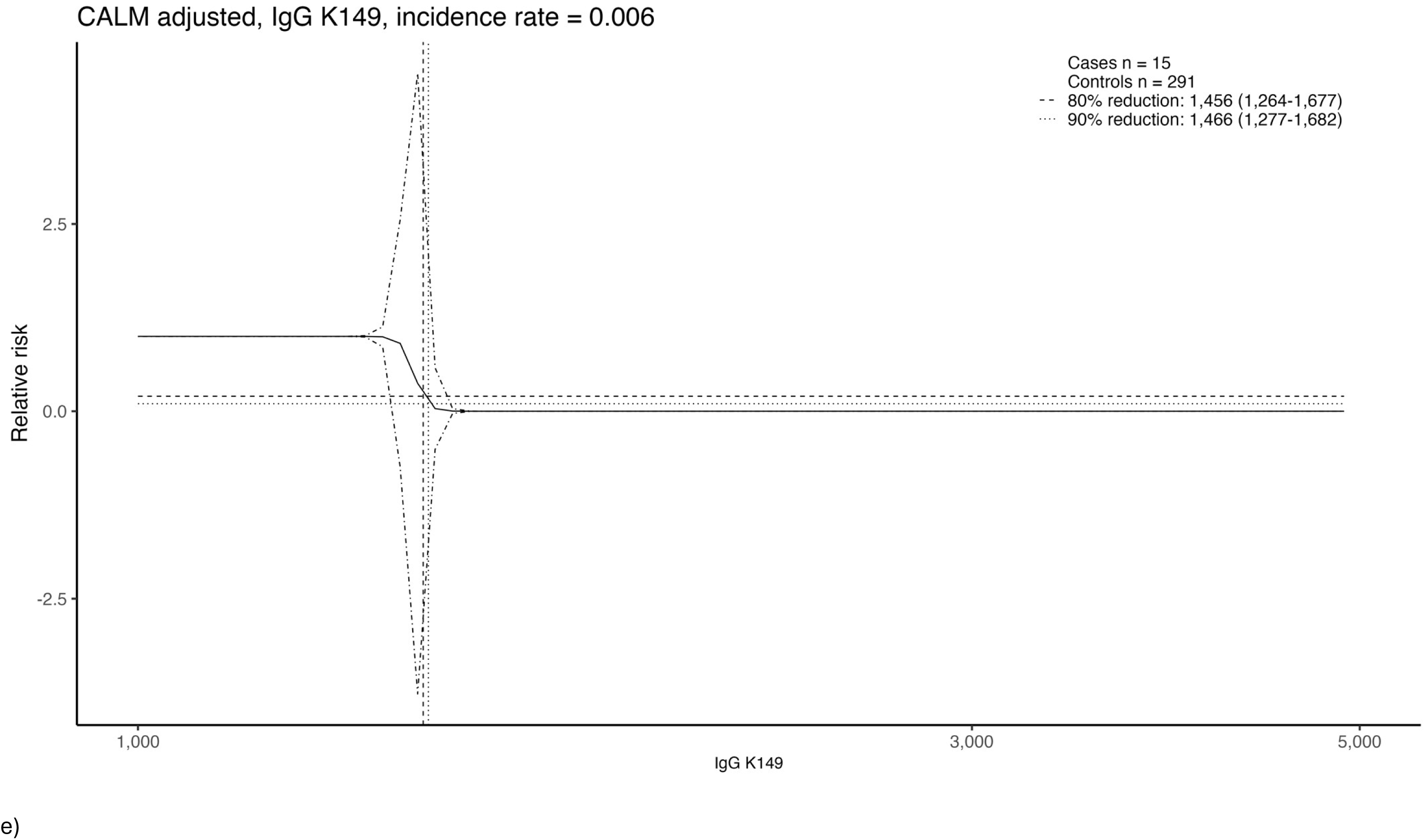
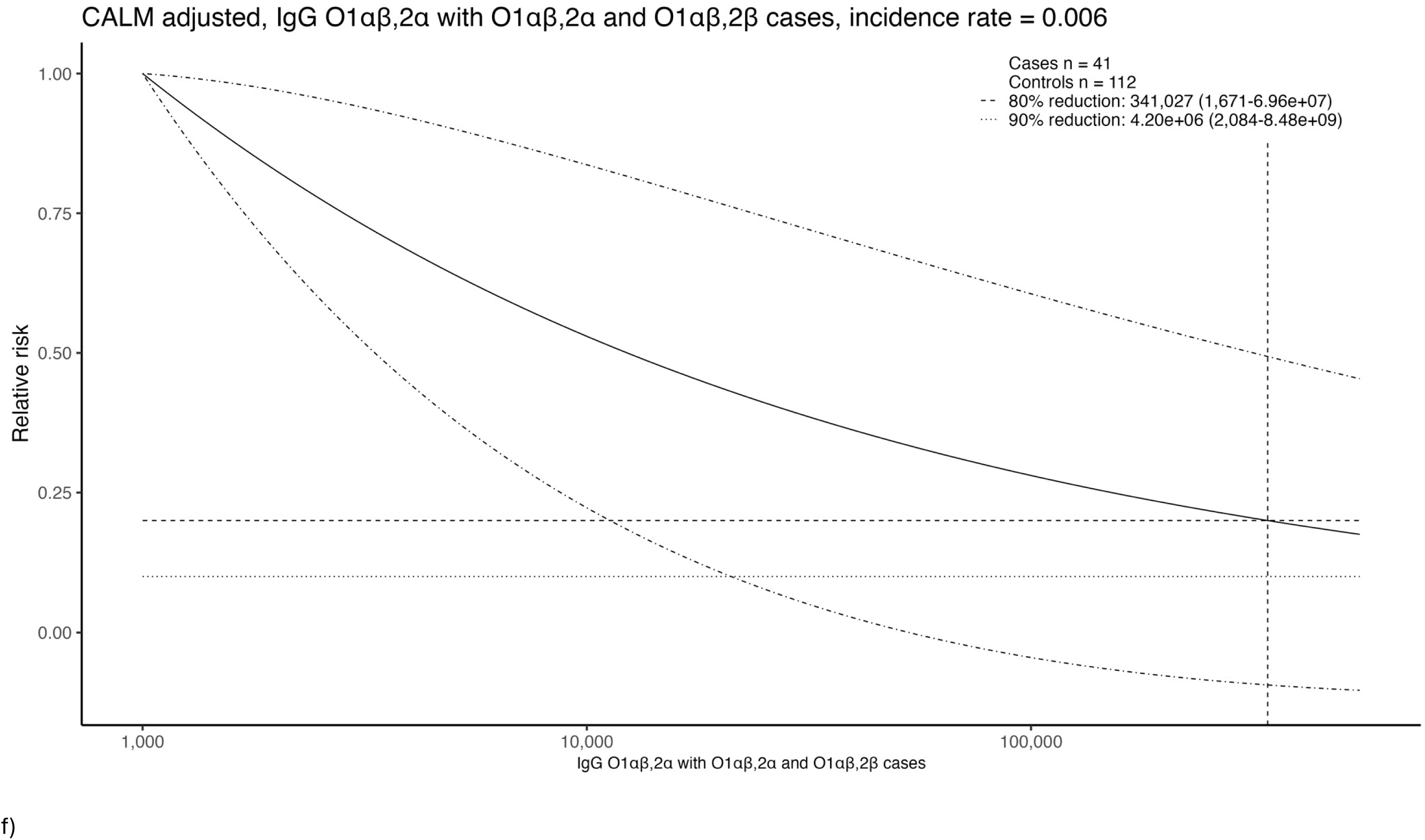
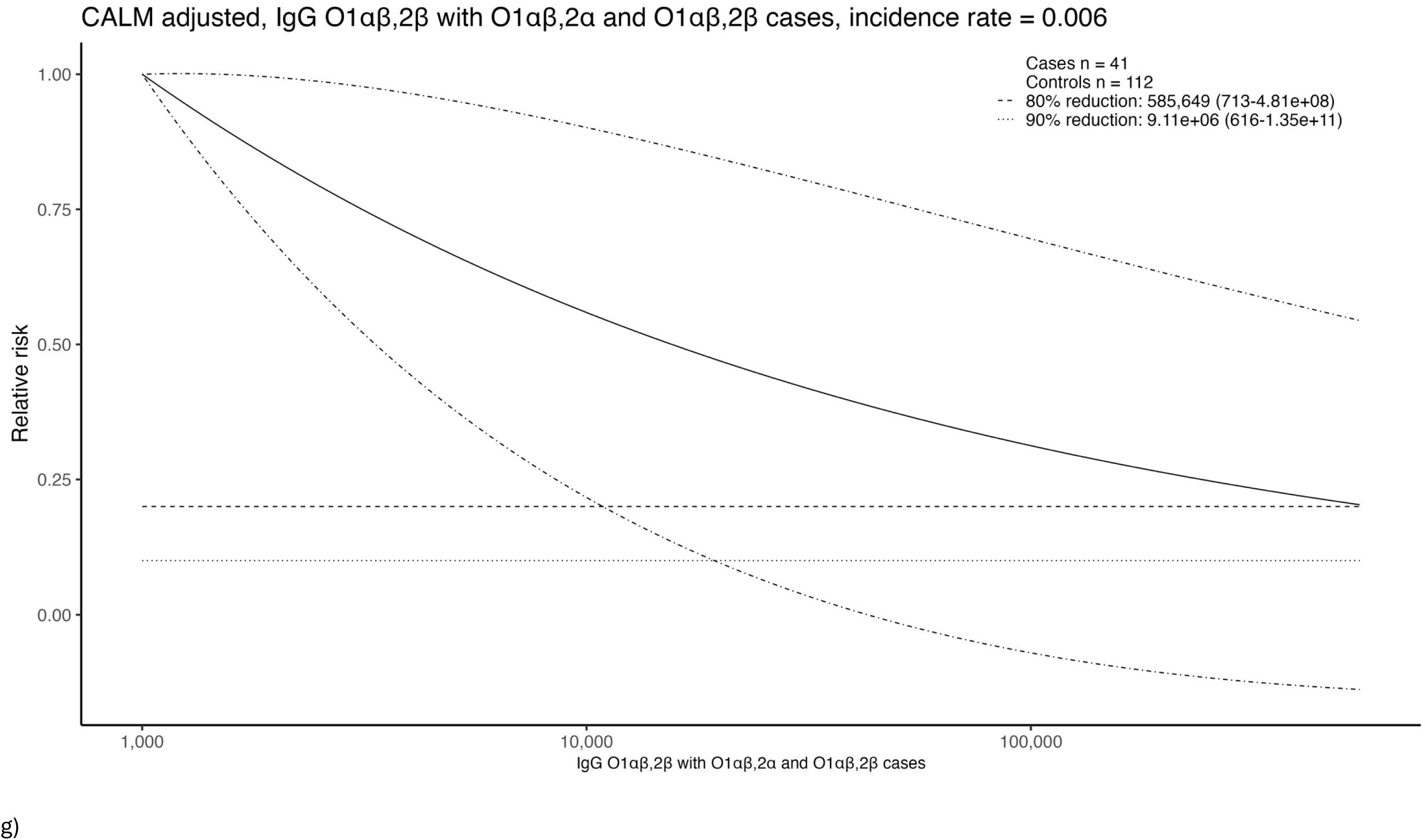
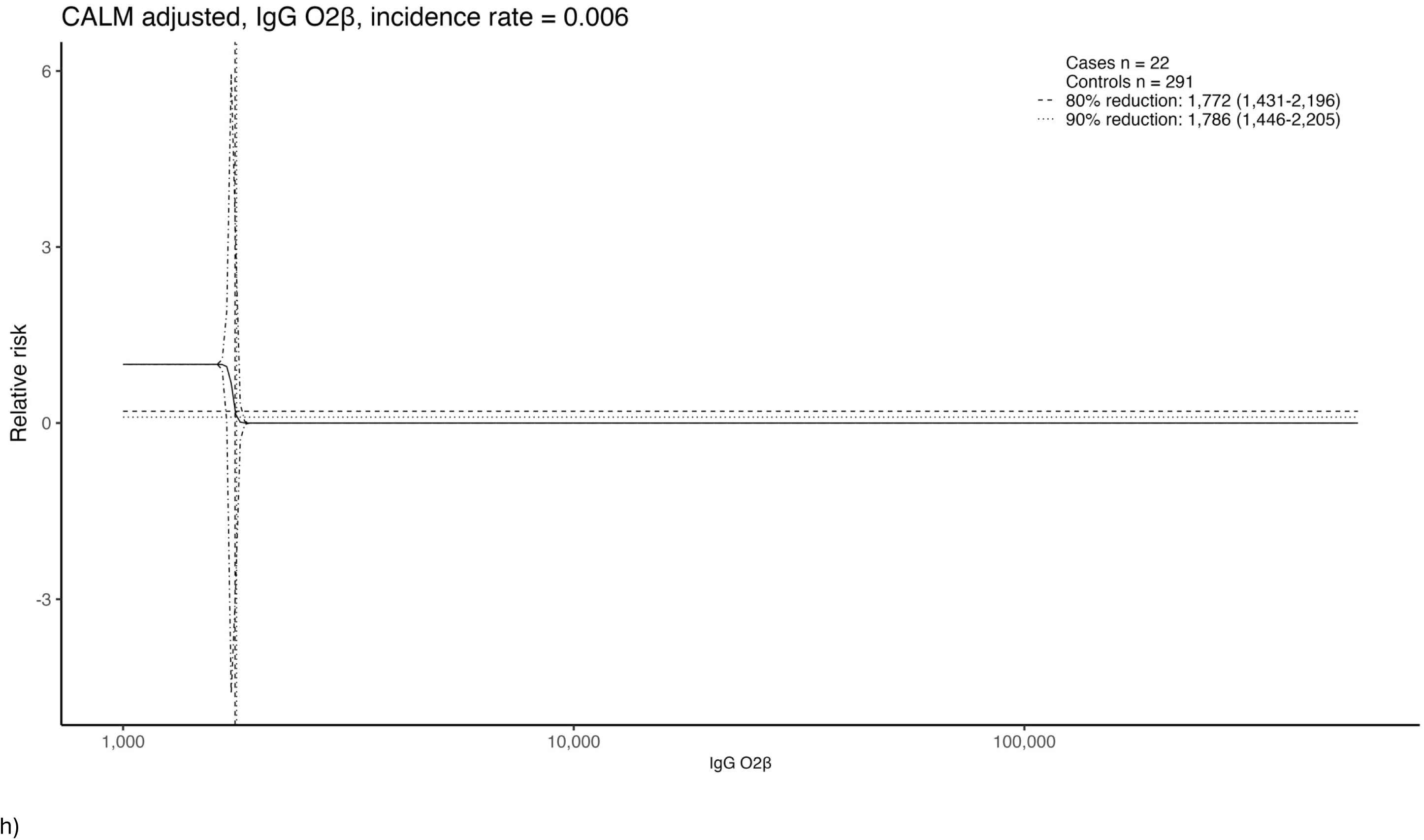
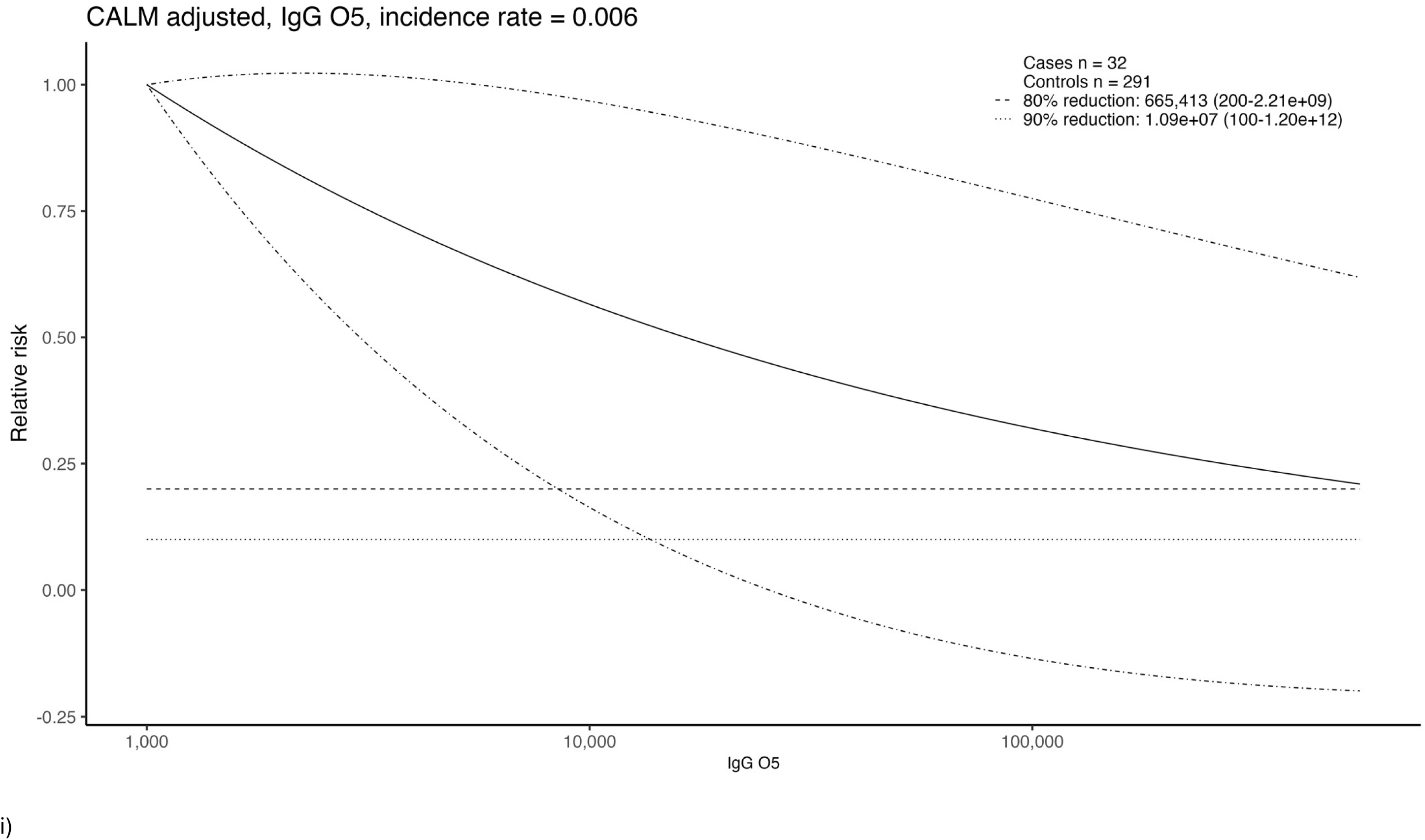
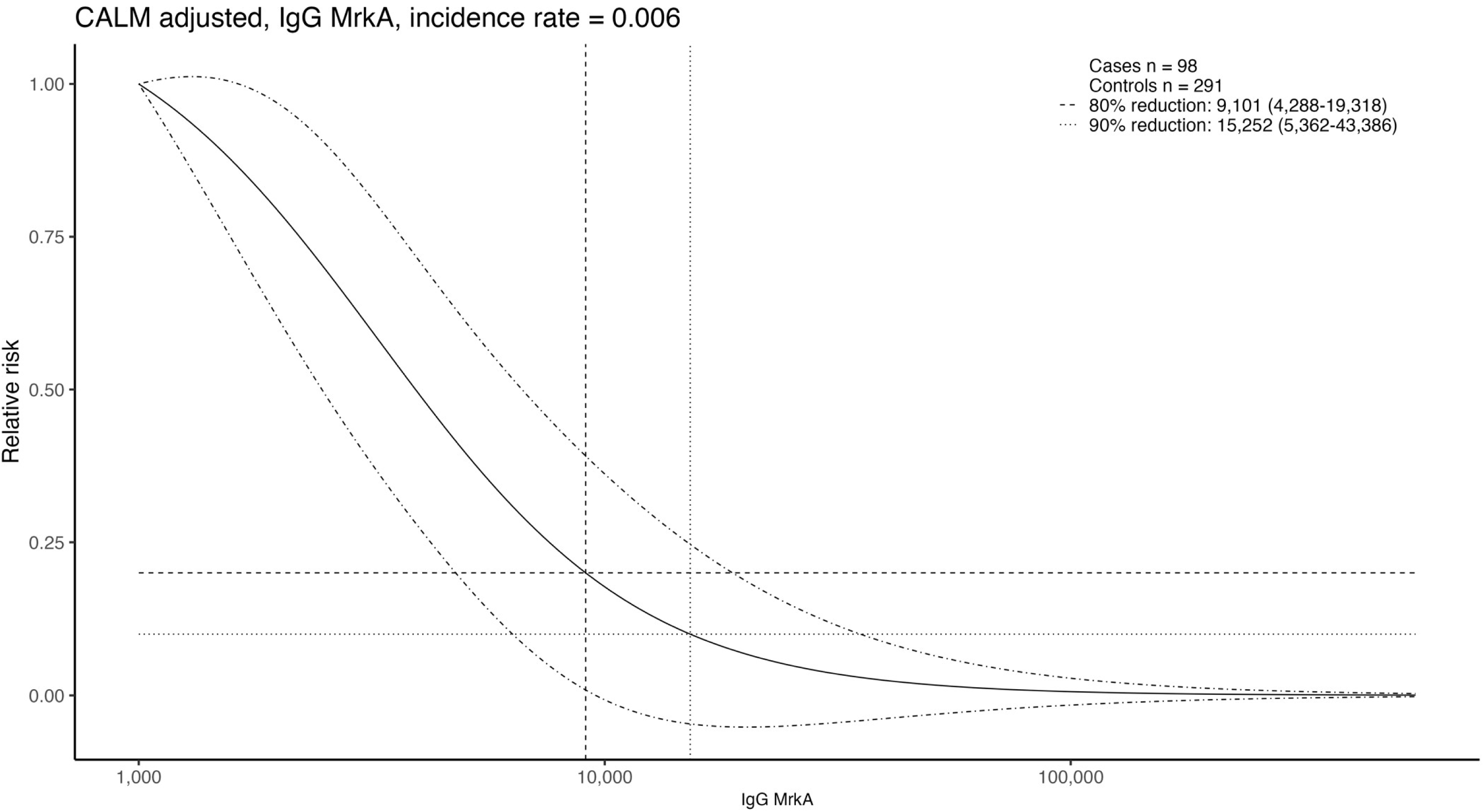
Estimated relative risk as a function of IgG for anti-K (a-d), anti-O (e-h), and MrkA IgG (i). IgG associated with a 50% and 80% reduction in disease is highlighted. Alt text: Graphical representation of estimated relative risk in disease for various anti-K, anti-O and MrkA IgG in subfigures a to i.

When comparing SBA GMTs between cases with the corresponding O-serotype of the SBA (Supplementary Table 1) and controls no differences were observed. Nevertheless, when comparing SBA GMTs between cases with the corresponding O-serotype and controls excluding values below LLoQ, O1αβ,2β cases had lower SBA GMTs (27 IC50; 95%CI: 12-63) compared with controls (136 IC50; 95% CI: 84-221; p=0.002; Table 4 and Figure 4b and Supplementary Figure 4b.

**Figure 4:**
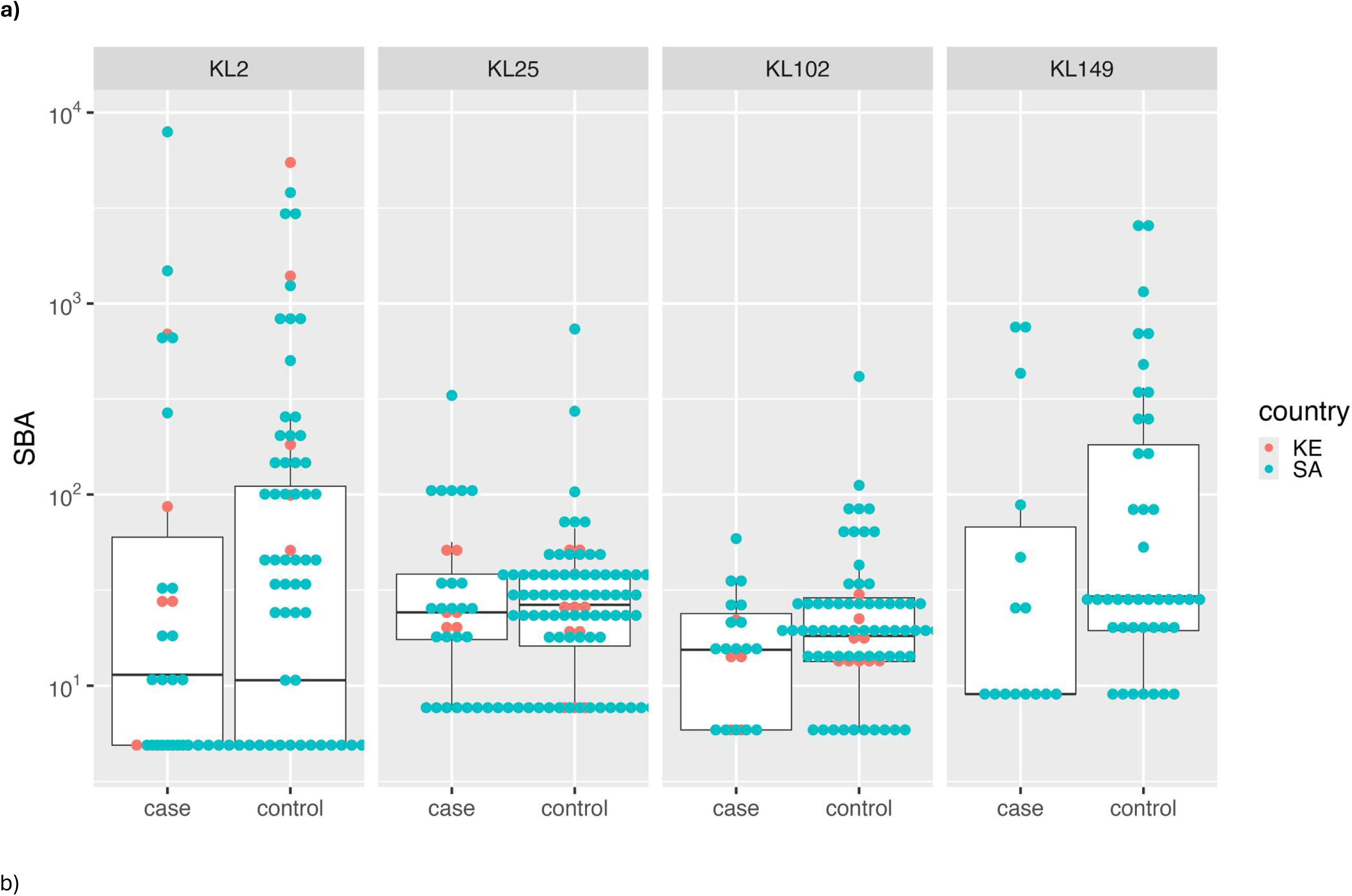
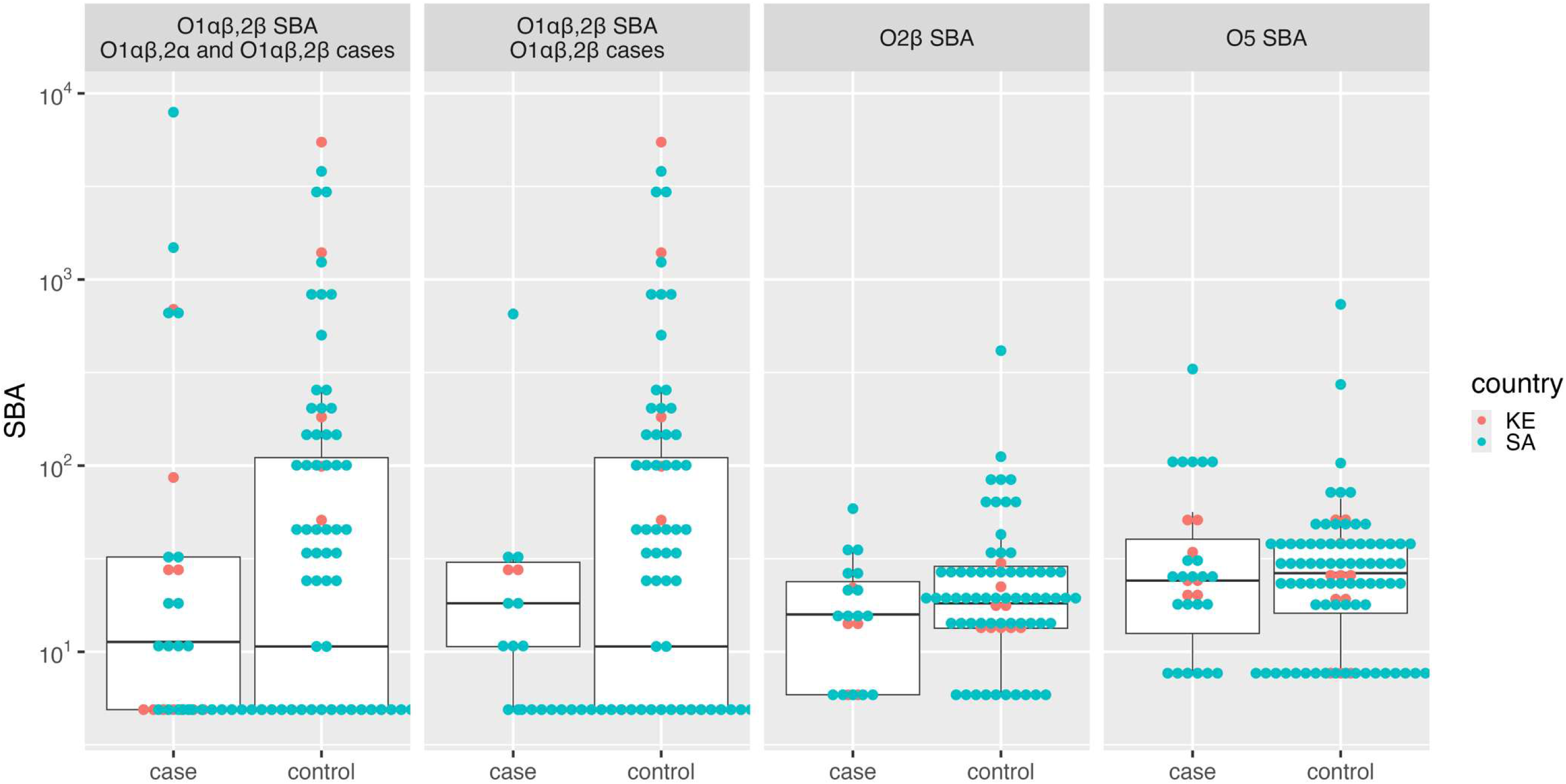
Dot plot with overlayed box-plots (whiskers extend to 1.5 times interquartile-range) of serum bactericidal assay values for K serotypes (a) and O serotypes (b).

Association between SBA and anti-O IgG was evaluated for O1αβ,2β, O2β and O5 in cord samples from infant controls; Supplementary Figure 5c. Correlation was moderate between O1αβ,2β SBA and O1αβ,2α IgG (Rho=0.643) as well as with O1αβ,2β IgG (Rho=0.657) and increased to 0.812 and 0.810 when excluding values below the LLoQ; Supplementary Figure 4d. Moderate correlation was observed between SBA and IgG for O5 (Rho=0.572) and low for O2β (correlation =0.048) and these estimates increased slightly when removing values below the LLoQ; Supplementary Figures 5c and 5d.

The GMC (RLU/mL) for anti-MrkA IgG were lower in cases (945; 95%CI: 757-1179) compared with controls (1607; 95%CI: 1378-1880; p<0.001); Table 3 and Figure 2c. This pattern was evident in a stratified analysis for all K- and O-serotypes, except for K149 and O4. The empirical distribution of cases and controls differs after 1500 RLU/mL; Supplementary Figure 3c. Minimal correlation was observed between SBA (combined assays) and MrkA IgG (Rho= 0.132 and 0.256 when removing values <LLoQ); Supplementary Figures 5e and 5f. There was substantial uncertainty in the estimated antibody concentration associated with an 80% reduction in iKPnD for MrkA IgG; Figure 3i and Supplementary Table 5.

A sensitivity analysis was conducted restricting controls to those matched to cases by birth weight, length of stay, and date of birth. The findings were consistent with those of the unmatched analysis; although significance was not maintained when comparing GMCs between cases and controls except for K2 and O1αβ,2α IgG; Supplementary Table 6. In a sensitivity analysis which was restricted to cord blood samples only, the results were similar to those obtained when both cord blood and acute serum samples were included; Supplementary Table 7.

## Discussion

In this exploratory study, we observed lower serotype-specific IgG GMCs to the homotypic K- or O-antigen in iKPnD cases for K2, K25 and O1αβ,2α types compared with controls without iKPnD. Furthermore, anti-MrkA IgG GMCs were also lower in cases across all anti-K and -O IgG, except for K25 and O4 compared with controls. Furthermore, we provide insight of the increasing efficiency of transplacental IgG transfer of K-, O- and MrkA antigens with increasing birth weight and gestational age; as well as geographic heterogeneity in efficiency of transplacental IgG transfer.

SToRR were observed for K, O-antigen and MrkA, suggesting immunity against iKPnD may be multifactorial, or that some markers of risk reduction are proxy markers rather than being mechanistically involved. Previous *K. pneumoniae* vaccine candidates include a multi-valent K-antigen polysaccharide formulation, and more recent efforts of an O-antigen-based vaccines.^12,30–33^ Our findings suggest a potential role of K antigens and some O antigens as vaccine targets. Also, if antibody against MrkA is indeed protective, its inclusion as a vaccine target with select K- and O-antigens could potentially enabling broader protection against multiple *K. pneumoniae* strains of diverse K- and O- serotypes. Differences in GMCs that were not observed for specific K and O-types in our study could become more evident with a larger sample size, which would provide greater power to detect differences. The data from our study also informs the sample sizes required to achieve adequate power to detect differences, which could guide in the design of future prospective studies investigating SToRR.

The delineation of SToRR could be a crucial step in the pathway to licensure of a *K. pneumoniae* vaccine for pregnant women, aimed at protecting their young infants. In our study, we observed infant IgG concentrations against K- and O- antigens were lower in cord blood samples from South African controls compared with those from Kenya, albeit the estimates being associated with wide confidence intervals. Additionally, CMR was higher in Kenya compared with in South Africa, which is consistent with previous observations that transplacental antibody transfer can vary geographically.

The sample size for the SBA analyses was smaller than for IgG because SBA measurements were only done against the homotypic antigen of the strain causing disease in the cases and their matched controls. Further studies integrating different immunological readouts, with larger samples size, and eventually contrasting different mechanism of functional responses will enable a better evaluation of the mechanism of protection. Notably, SBA correlated strongly with anti-O IgG responses, with correlation further strengthened when analyses were restricted to samples above the LLoQ for both assays. By contrast, correlations between SBA and anti-MrkA or anti-K IgG were weak.

Our study has several limitations, including that the exposure status of control participants was not directly measured, which could bias estimates of association. Under the assumption that hospital exposure is a prerequisite for invasive disease, control participants who were not exposed to KL2, KL25, KL102, or KL149 *K. pneumoniae* serotypes would not have been at risk of developing disease, potentially leading to biased effect estimates. Nevertheless, we estimate that approximately at least 70% of hospitalized children experience exposure to *K. pneumoniae*, which may partially mitigate this concern.^24^ The mechanism of natural immunity within the general population is unknown and immune factors beyond those evaluated in this analysis may contribute to protection against disease. Control participants had been initially selected to be matched to cases on birth weight, length of hospital stay, and age (within 60 days). Although the final analysis incorporated all available controls rather than maintaining the matched design, the initial control selection process may have introduced bias into effect estimates. Nevertheless, birth weight and country were adjusted for in the analytical models to mitigate potential bias. Additionally, as a sensitivity analysis, we repeated the analysis using matched cases and controls and found that the results were largely consistent with the unmatched analysis.

Although exploratory and despite the above limitations, our study provides the first measurements of immune markers against potential vaccine targets in relation to risk of iKPnD in young infants, the findings of which could inform future studies, including simulation-based analyses to estimate the sample sizes required to detect meaningful differences to characterise SToRR for iKPnD. Future work to incorporate systems serology approaches to look at potential other Fc Receptor mediated activity of antibodies, their avidity, as well as contrasting different functional readouts is also underway and will enable a more comprehensive characterization of immune responses and provide deeper insight into correlates of natural immunity.

## Author contributions

SM, JAB, NB, UNN, FM, OR and ZD conceptualised the study design and defined the objectives of the manuscript. ZD, ND supervised data collection in South Africa, and AA, MN, AN, JAB supervised data collection in Kenya. MC, LR, GFB and OR performed all laboratory assays. AI developed and implemented the statistical analysis plan. AI and SM drafted the initial version of the manuscript. FM, OR, UNN provided critical review and substantive revisions to the manuscript. All authors critically reviewed and approved the final manuscript.

## Acknowledgments

The authors thank the study participants for their willingness to take part in this research. We also acknowledge the WITS-VIDA and KWTRP study teams for their valuable contributions to data collection and data management as well as the Gates Foundation for funding this work.

## Data availability

De-identified data used in this analysis available upon request from WITS-VIDA on a collaborative basis provided a data transfer agreement is approved. All requests for data sharing will require approval of WITS-VIDA, data governance committee, KWTRP and GSK Biologicals SA.

