## Supplementary for "Association of serum antibody to serotype-specific capsular (K), lipopolysaccharide (O) and MrkA with risk reduction of invasive *Klebsiella pneumoniae* disease in young infants: an observational study"

### Supplementary material

#### Methods

*Study design*
In South Africa, iKPnD cases were enrolled from two previous cohort studies designed to evaluate serological thresholds of risk reduction (SToRR) against invasive GBS disease.[1,2] Briefly, mother-newborn dyads were enrolled at the time of birth at two hospitals in Johannesburg, South Africa, and maternal and cord blood samples were collected. The blood samples were centrifuged within 12 hours of collection at the Wits Vaccines and Infectious Diseases Analytics Unit (Wits-VIDA), and serum was aliquoted and archived at -70 °C. All the newborns were followed up with hospital-based surveillance for hospitalization through 90 days of age. Medical records and laboratory results were abstracted for all cohort infants who were hospitalised. In parallel, we also enrolled iKPnD cases from the same two hospitals with culture-confirmed iKPnD who were not part of the cohort (i.e., non-cohort cases). For the non-cohort cases, venous blood samples were obtained within 72 hours of laboratory confirmation of iKPnD and processed at Wits-VIDA. The controls in South Africa were selected from infants enrolled in the cohorts who met the following criteria: (i) maternal and cord blood available, and (ii) they were admitted to hospital with no suspected sepsis during hospitalisation.

Blood samples for bacterial culture were collected aseptically by venepuncture and inoculated into paediatric blood culture bottles (bioMérieux, South Africa). Cultures were processed by the National Health Laboratory Service (NHLS) according to standard operating procedures and incubated using the BacT/ALERT® automated blood culture system (bioMérieux, Marcy l'Etoile, France). Alarm-positive blood cultures underwent Gram staining and subculture for organism isolation. Cerebrospinal fluid specimens were cultured using standard microbiological methods. Bacterial identification and antimicrobial susceptibility testing were performed using conventional techniques and automated systems, including VITEK® 2 (bioMérieux). The study team collected the bacterial isolates cultured on blood or CSF from NHLS, including *K. pneumoniae* isolates, which were archived at Wits-VIDA. Upon receipt, isolates were subcultured onto Chromogenic Colorex™ Orientation agar to confirm viability, purity, and organism identity. Confirmed *K. pneumoniae* isolates were inoculated into skim milk–tryptone–glucose–glycerol (STGG) medium and stored at −70 °C until whole-genome sequencing (WGS) done at the National Institute for Communicable Disease, Johannesburg, South Africa.

In Kenya, blood and CSF cultures were transported to KEMRI Wellcome Trust Research Programme (KWTRP) laboratories. Blood cultures were incubated in BACTEC incubators (BD, USA) and positive isolates were identified by API (bioMérieux, France) up to 2018 and thereafter by matrix assisted laser desorption ionization – time of flight (MALDI-TOF, Bruker, Germany). CSF samples were cultured using standard microbiological techniques. Whole genome sequencing was done as previously described.[3] Briefly, DNA was extracted using QIAamp Fast DNA stool mini kit, libraries were prepared using the Illumina DNA prep kit and sequencing was done on the MiSeq platform (Illumina, USA) in Kilifi. Quality control was performed using FastQC v0.12.1 before de novo assembly with SPAdes v3.15.5. Capsule and O - antigen predictions was performed using Kaptive within Kleborate v3.2.4 pipeline

A maternal surveillance (KIPMAT) to detect serious adverse events in pregnancy and inform clinical care was conducted in Kilifi from 2011 to 2020 and cord blood and maternal blood samples were collected during delivery.[4] Neonatal babies born during the KIPMAT surveillance and who were admitted had a blood sample collected for culture and clinical chemistry and part of this sample was stored.

*Laboratory methods*
Details of the multiplex bead-based assay and serum bactericidal assay are described below.

*Genomic sequencing*
Genomic DNA was extracted from STGG stocks using the Whitesci Tanbead® Nucleic Acid Extraction System Maelstrom 4800 (Taiwan Advanced Nanotech) following the manufacturer's recommendations. Libraries were prepared using Nextera XT and sequenced on the Illumina NextSeq platform (150 bp paired-end) s,. Sequence data were processed using the Jekesa bioinformatics pipeline for bacterial pathogens.[2] Quality control included removal of reads with Phred scores <30 and read lengths <50 bp, and contamination screening using ConFindr. De novo genome assemblies were generated using SPAdes and Shovill and assessed with QUAST, followed by annotation using the RAST server. Multilocus sequence typing (MLST) and core genome MLST (cgMLST) were performed using the PasteurMLST and BIGSdb-Pasteur platforms, respectively. Capsular and O-antigen serotypes were predicted using the Kleborate pipeline against the Kaptive v3 database. Antimicrobial resistance genes, virulence factors, and plasmid replicons were identified using ResFinder, VirulenceFinder, PlasmidFinder v2.1, and Kleborate, incorporating CARD and ResFinder v9 databases.

*Multiplex bead-based assay for IgG determination*
For binding assays polysaccharidic biotinylated antigens or MrkA were loaded to streptavinitaded beads of different region at a concentration of 1 µg/mL and 15 µg/mL of each antigen, respectively. A total of 50 µL of diluted standard or sample sera was mixed with 10 µL of containing 2500 beads/region/well in a 96-well Greiner plate (655904; Greiner Bio-One, Kremsmünster, Austria). Plates were incubated for 60 min at room temperature in the dark on a plate shaker at 750 rpm and afterwards washed three times with 200 µL of PBS. Each well was loaded with 50 µL of 10 µg/mL R-phycoerythrin AffiniPure goat anti-human IgG, Fcγ-fragment-specific (Jackson Immunoresearch code 109-116-098) in PBS, then incubated for 60 min in the dark under shaking at 750 rpm. After incubation, beads were washed three times with 200 µL of PBS, then resuspended in 100 µL of PBS and acquired using a Bioplex-200 reader, reading at least 50 beads per region at a high RP1 target. An 11-point, 3-fold serially diluted calibrated standard serum prepared in 10 mM PBS pH 7.2 was run in duplicate for all plates together with two blank wells containing PBS only were included in each plate. Test serum samples were prepared at 4 dilution points in 3-fold serial dilutions. A 5PL parameter logistic curve was fit to the blank-subtracted median fluorescence

intensity (MFI) values obtained for each standard curve point using Bioplex Manager 6.2 Software (Biorad, Watford, UK). The Relative Luminex Units/mL (RLU/mL) for each dilution of samples were obtained by interpolation of the blank subtracted MFI values at each specific dilution against the 5PL parameter standard logistic curve multiplied for the initial dilution. Only values falling within the dynamic range of the fitted standard curve were considered for the analysis (automatically kept by Bioplex manager Version 6.2, Biorad, Watford, UK). Interpolation of the standard curves allowed the blank-subtracted MFI values for samples to be converted to RLU/mL. Several QC criteria were applied both to the standard and the sample to validate the run or the single measurement, respectively. The plate was considered valid if the mean of the %relative error of the observed concentration of each calibrator of the standard curve calculated respective of the nominal concentration of the calibrator was equal or less then 20% and if the RLU/mL of two control samples fell within the expected range. The concentration in RLU/mL for each sample was calculated as the geomean of the values of the dilutions (at least two) tested for which the %relative error of each dilution versus the median of the RLU/mL of the four dilutions was within the 30%. The sample was considered valid if the %CV of valid geomeans were equal or less then 30%. Results of the assay are Relative Luminex Units/mL, with 1 RLU/mL defined as the reciprocal of the sera dilution that gave 1000 MFI in a standard assay.

*Luminescent based Serum Bactericidal Assay to determine sera functionality.*
Different dilutions of test sera (heat inactivated – HI - at 56°C for 30 min) were incubated with bacteria (approximately 10,000 bacteria/well) in 96-well round bottom sterile plates (Corning, Glendale, AZ, USA in the presence of hexogenous Baby Rabbit Complement (BRC) at a final concentration of 50% for K2, 20% for K25 and 30% for K102 and K149. . The HI sera were serially diluted in LB in the SBA plate (25 µL/well) starting from 1:10 dilution (final dilution) followed by 3-fold dilution steps up to 7 dilution points plus one control well with no sera. Log-phase cultures for the assay at OD600 of 0.22 ± 0.03 were prepared by starting a fresh culture from OD600 = 0.05 from an overnight (16–18 h) culture of bacteria grown in LB at 37°C stirring from a frozen −80 °C bacterial working aliquot. Final reaction volume of the mix was 100 µL/well and plate were incubated for 3 h at 37 °C. At the end of the incubation, the SBA plate was centrifuged at room temperature for 10 min at 4000 × g, the supernatant was discarded the bacterial pellets were resuspended in PBS and transferred to a white 96-well plate (Greiner Bio-One, Roma, Italy) and mixed 1:1 v:v with BacTiter-Glo Reagent (Promega, Southampton, UK). The reaction was incubated for 5 min at room temperature on an orbital shaker at 600 rpm, and the luminescence signal was measured by a luminometer (Synergy HT, Biotek, Swindon, UK). A 4-parameter non-linear regression was applied to the raw luminescence obtained for all the sera dilutions tested assigning an arbitrary serum dilution of $10^{15}$ to the well containing no sera. Fitting was performed by weighting the data for the inverse of luminescence. To validate the dilution series, the highest luminescence detected in the dilution series at T180 had to be at least 0.7-fold the luminescence detected in the control well with no sera and the IC50 for the standard sera, run on each plate, within the predefined acceptable range.

*Statistical Methods*

The analysis plan specified that the association between birth weight (used as a proxy for GA) and serotype-specific IgG in cord blood samples of infant controls would be evaluated to inform whether a matched or unmatched population would be used for estimating the association between immunological measurements of natural immunity and iKPnD. If no association was observed between birth weight and serotype-specific IgG in the controls then the matched population would serve as the primary analysis set, as the matching would not bias the estimates. Conversely. if a moderate to strong correlation was observed between birth weight and IgG, as was the case in the study, the unmatched population was used as the primary analysis set. As a sensitivity analysis, the analyses were repeated using the matched population.

A CALM model was fit to anti-K, anti-O and MrkA infant IgG in cases and controls adjusting for low-birth weight (1500g-<2500g), very-low birth weight (<1500g) and country. Briefly, the CALM allows for incorporation of covariates when estimating the relationship between assay values and disease, assuming that covariates do not modify this relationship. Under this assumption, the probability of disease is estimated by a product of the probability of disease given covariates and the risk reduction of disease associated with assay value relative to the probability of disease at the lowest assay value. The second term of this product corresponds to the $RRR_a$. Inverse probability weighting was employed to adjust for the case-control design.

ETHICS

The collection of samples from the cohort studies in South Africa, as well as the enrolment of the non-cohort cases, was approved by the University of the Witwatersrand Human Ethics Research Committee (Wits HREC reference: 181110 and Wits HREC reference: 220214). Signed informed consent from parents included permission for additional use of blood samples beyond those stipulated in the study protocol, subject to further Ethics approval. The testing of samples for this study was approved separately by Wits HREC (Ref: M230652). In Kenya, ethical approval to conduct the surveillance studies was provided by KEMRI Scientific and Ethic Review (SERU) and written informed consent was obtained from the participant or guardian (SCC1433 for bacteraemia surveillance and SCC1778 for KIPMAT study). Approval for further analysis for this study was provided by SERU (SERU 4687).

### Results

*Maternal SBA and cord-maternal ratio of SBA with birth weight and GA*

The CMRs for SBA were also modelled by GA and BW. However, 35% (93/269) of available paired samples had values from either maternal or cord blood below the LLOQ and for which CMR was not estimable (Supplementary Table 2). Of the remaining 176 samples, only 5 were from Kenya and consequently the analysis was restricted to South African samples. Modelled estimates of CMR rose with increasing birth weight and GA for K2 and K25, whereas, CMR SBA decreased for K102 and K149 as birth weight and GA increased. (Supplementary Figure 2). Modelled CMR for IgG was below 0.5 for GA of 20 weeks and increased progressively to approximately 0.75-1.0 by 42 weeks GA across all K-types (Figure 1b). In contrast, modelled CMR for SBA against K102 and K149 remained consistently below 0.5 across the GA range (Supplementary Figure 2b).

For K2, the modelled CMR for SBA was close to zero at 20 weeks GA and increased to values exceeding 2 beyond 38 weeks GA. However, this apparent increase was driven by two influential observations from infants born at 39 weeks GA, with CMR values of 13.1 and 191.5. This pattern was not observed in analyses stratified by birth weight, as the corresponding birth weights were 1,765 g and 3,410 g, respectively. For SBA against K25, modelled CMR estimates remained stable, ranging between 0.5 and 1.0 across all gestational ages.

**Supplementary Table and Figures**

**Supplementary  Table 1: Unit and lower limit of quantification for laboratory tests measuring IgG and SBA.**

| Method | Test | Unit | LLOQ[1] |
|---|---|---|---|
| Luminex | IgG MrkA | RLU/mL | 4 |
| Luminex | IgG K2 | RLU/mL | 9 |
| Luminex | IgG K25 | RLU/mL | 6 |
| Luminex | IgG K102 | RLU/mL | 11 |
| Luminex | IgG K149 | RLU/mL | 12 |
| Luminex | IgG O1αβ,2α | RLU/mL | 27 |
| Luminex | IgG O1αβ,2β | RLU/mL | 17 |
| Luminex | IgG O2α | RLU/mL | 10 |
| Luminex | IgG O2β | RLU/mL | 18 |
| Luminex | IgG O5 | RLU/mL | 4 |
| SBA | K2; O1αβ,2β | IC50 | 10 |
| SBA | K25; O5 | IC50 | 15 |
| SBA | K102; O2β | IC50 | 12 |
| SBA | K149; O4 | IC50 | 18 |

[1]LLOQ: Lower limit of quantification

**Supplementary  Table 2: Summary of analyses.**

| Objective | Methods | Analysis subset |
|---|---|---|
| Correlation between serotype-specific IgG and birth weight | Simple linear regression with serotype-specific IgG as dependent variable and birth weight as independent variable | Cord blood samples from South African and Kenyan infant controls.<br>280 samples (30 KE, 250 SA) |
| Correlation between serotype-specific IgG and gestational age | Simple linear regression with serotype-specific IgG as dependent variable and gestational age as independent variable | Cord blood samples from South African infant controls.<br>180 samples[1] |
| Association of cord:maternal ratio (CMR) of serotype-specific IgG and birth weight | Simple linear regression with serotype-specific CMR as dependent variable and gestational age as independent variable | Cord and maternal blood paired samples from South African and Kenyan infant controls.<br>270[2] paired samples (20 KE, 250 SA) |
| Association of cord:maternal ratio (CMR) of serotype-specific IgG and gestational age | Simple linear regression with serotype-specific CMR as dependent variable and gestational age as independent variable | Cord and maternal blood paired samples from South African infant controls.<br>180 samples[1,2] |
| Association of cord:maternal ratio (CMR) of SBA and birth weight | Simple linear regression with SBA CMR as dependent variable and gestational age as independent variable | Cord and maternal blood paired from South African infant controls.<br>269[3] (20 KE, 249 SA) tested paired samples (176 both samples >LLOQ).<br>K2: 77 (10 KE, 67 SA); both samples >LLOQ: 29 (1 KE, 28 SA)<br>K25: 81 (40 KE, 77 SA); both samples >LLOQ: 55 (0 KE, 55 SA)<br>K102: 69 (6 KE, 63 SA) ; both samples >LLOQ: 57 (4 KE, 53 SA)<br>K149: 42 (0 KE, 42 SA); both samples >LLOQ: 35 (0 KE, 35 SA) |
| Association of cord:maternal ratio (CMR) of SBA and gestational age | Simple linear regression with SBA CMR as dependent variable and gestational age as independent variable | Cord and maternal blood paired from South African infant controls<br>180 tested paired samples (119 both samples >LLOQ).<br>K2: 47 (17 both samples >LLOQ)<br>K25: 42 (28 both samples >LLOQ)<br>K102: 55 (45 both samples >LLOQ)<br>K149: 36 (29 both samples >LLOQ) |
| Association between serotype-specific IgG IKPnD | Empirical distribution of IgG in both case and controls<br>GMCs (and 95% Cis) of IgG in both cases and controls<br>CALM with IKPnD as dependent variable and IgG, LBW, VLBW and country as covariates. | Cord and acute blood samples from South African and Kenyan infant cases and controls.<br>Breakdown of available samples is detailed in Table 1.<br>*Sensitivity analysis*: cord blood samples from South African and Kenyan infants in cases and controls |
| Association between SBA and IKPnD | Empirical distribution of SBA in both case and controls<br>GMTs (and 95% CIs) of SBA in both cases and controls<br>CALM with IKPnD se as dependent variable and SBA, LBW, VLBW and country as covariates. | Cord and acute blood samples from South African and Kenyan infant cases and controls<br>Breakdown of tested samples is detailed in Table 1.<br>*Sensitivity analysis:* cord blood samples from South African and Kenyan infants in cases and controls |

| | | |
|---|---|---|
| Correlation between SBA and serotype-specific IgG | Pearson's correlation coefficient<br>Scatterplot of SBA and serotype specific IgG | Cord blood samples from South African and Kenyan infant controls<br><br>K2: 84 (43 both IgG and SBA >LLOQ); O1αβ,2β: 41 (25 both IgG and SBA >LLOQ)<br>K25: 82 (62 both IgG and SBA >LLOQ); O5: 81 (61 both IgG and SBA >LLOQ)<br>K102: 72 (59 both IgG and SBA >LLOQ); O2β: 68 (56 both IgG and SBA >LLOQ)<br>K149: 42 (35 both IgG and SBA >LLOQ)<br><br>*Sensitivity analysis:* Cord blood samples from South African and Kenyan infant controls where assay value is above the LLOQ |

[1]70 of the 250 SA samples were missing gestational age; [2]10 Kenyan controls did not have maternal blood. 1 SA sample had both infant and maternal blood <LLOQ for anti-K25 IgG and maternal blood <LLOQ for anti-O2v2 IgG. [3]10 Kenyan controls did not have maternal blood and one SA sample had SBA K2 performed on the cord blood and SBA K25 on maternal blood.

**Supplementary Table 3**: Parameter estimates (and standard errors) from a simple linear regression of log cord IgG from infant controls adjusting for birth weight and country or gestational age.

| | K2 | K25 | K102 | K149 | O1αβ,2α | O1αβ,2β | O2α | O2β | O5 | MrkA |
|---|---|---|---|---|---|---|---|---|---|---|
| Coefficient for birth weight (g) | 0.19 (0.11) | 0.27 (0.11) | 0.27 (0.09) | 0.22 (0.08) | 0.13 (0.11) | 0.15 (0.11) | 0.3 (0.09) | 0.12 (0.08) | 0.17 (0.09) | 0.29 (0.1) |
| Coefficient for country | -0.07 (0.27) | -0.78 (0.28) | -0.42 (0.24) | -0.1 (0.21) | 0.29 (0.29) | 0.31 (0.28) | 0.15 (0.24) | 0.21 (0.19) | 0.1 (0.23) | 0.11 (0.26) |
| Coefficient for GA | 0.03 (0.02) | 0.03 (0.02) | 0.05 (0.02) | 0.04 (0.02) | 0.02 (0.02) | 0.03 (0.02) | 0.06 (0.02) | 0.02 (0.02) | 0.05 (0.02) | 0.05 (0.02) |

**Supplementary Table 4:** Parameter estimates (and standard errors) from a simple linear regression of log cord:maternal ratio of IgG from infant controls adjusting for birth weight and country or gestational age.

| | K2 | K25 | K102 | K149 | O1αβ,2α | O1αβ,2β | O2α | O2β | O5 | MrkA |
|---|---|---|---|---|---|---|---|---|---|---|
| Coefficient for birth weight (g) | 0.28 (0.06) | 0.24 (0.07) | 0.3 (0.07) | 0.19 (0.06) | 0.27 (0.07) | 0.28 (0.07) | 0.29 (0.06) | 0.16 (0.07) | 0.22 (0.07) | 0.28 (0.06) |
| Coefficient for country | -0.14 (0.19) | -0.19 (0.2) | -0.1 (0.2) | -0.22 (0.19) | -0.22 (0.22) | -0.32 (0.22) | -0.18 (0.19) | -0.18 (0.2) | -0.25 (0.21) | -0.11 (0.18) |
| Coefficient for GA | 0.04 (0.01) | 0.03 (0.01) | 0.05 (0.01) | 0.03 (0.01) | 0.03 (0.01) | 0.03 (0.01) | 0.05 (0.01) | 0.02 (0.01) | 0.03 (0.01) | 0.04 (0.01) |

**Supplementary Table 5: Antibody concentrations associated with an 50% and 90% estimated risk reduction using CALM model adjusting for low birth weight, very low birth weight and country.**

| Case definition | IgG | Number of cases | Number of controls[1] | 50% reduction | | 80% reduction | |
|---|---|---|---|---|---|---|---|
| | | | | IgG | 95%CI | IgG | 95%CI |
| K2 | K2 | 28 | 291 | 4,680 | 586 to 37,352 | 22,396 | 240 to >100,000 |
| K25 | K25 | 33 | 291 | 1,980 | 1,326 to 2,957 | 4,333 | 1,763 to 10,651 |
| K102 | K102 | 22 | 291 | 34,569 | 0 to >100,000 | - | - |
| K149[2] | K149 | 15 | 291 | 1,439 | 1,240 to 1,670 | 1,456 | 1,264 to 1,677 |
| O1αβ,2α | O1αβ,2α | 16 | 291 | 13,048 | 227 to >100,000 | - | - |
| O1αβ,2β | O1αβ,2β | 25 | 291 | 246 | 131 to 462 | 243 | 142 to 417 |
| O1αβ, 2α or O1αβ, 2β | O1αβ,2α | 41 | 112 | 12,326 | 1,247 to >100,000 | - | - |
| O1αβ, 2α or O1αβ, 2β | O1αβ,2β | 41 | 112 | 15,558 | 864 to >100,000 | - | - |
| O2β | O2β | 22 | 291 | 1,750 | 1,404 to 2,181 | 1,772 | 1,431 to 2,196 |
| O5 | O5 | 32 | 291 | 16,438 | 500 to >100,000 | - | - |
| All cases | MrkA | 98 | 291 | 3,926 | 2,381 to 6,473 | 9,101 | 4,288 to 19,318 |

[1]1 control from Kenya had unknown birth weight and was excluded in the model. [2]Cases for K149 were only from South Africa and there were no cases with normal birth weight, thus only very low birth weight was included as a covariate in the CALM.

**Supplementary Table 6**: **Geometric mean concentrations (and 95% confidence intervals) between cord blood or acute serum[1] from infants with invasive *Klebsiella pneumoniae* and those without disease which were matched to cases for anti-K and anti-O IgG antibodies.**

| Case | IgG | Case [RLU/mL] | Control [RLU/mL] | p-value |
|---|---|---|---|---|
| K2 | K2 | 396 (250-628) n=28 | 637 (493-822) n=83 | 0.074 |
| K25 | K25 | 396 (251-623) n=33 | 1048 (786-1396) n=87 | 0.001 |
| K102 | K102 | 503 (295-859) n=22 | 602 (447-809) n=73 | 0.553 |
| K149 | K149 | 327 (204-521) n=15 | 479 (307-748) n=33 | 0.224 |
| O1αβ,2α | O1αβ,2α | 1224 (436-3434) n=16 | 1976 (1271-3072) n=37 | 0.379 |
| O1αβ,2β | O1αβ,2β | 1067 (642-1774) n=25 | 1978 (1362-2873) n=76 | 0.052 |

| | | | | |
|---|---|---|---|---|
| O1αβ,2α or O1αβ,2β[2] | O1αβ,2α | 1282 (782-2101) n=41 | 2393 (1786-3206) n=113 | 0.032 |
| O1αβ,2α or O1αβ,2β[2] | O1αβ,2β | 1059 (661-1697) n=41 | 1828 (1375-2430) n=113 | 0.050 |
| O2β | O2β | 358 (255-503) n=22 | 431 (338-549) n=73 | 0.369 |
| O5 | O5 | 695 (456-1058) n=32 | 1069 (825-1384) n=86 | 0.083 |

[1]Acute serum is the serum obtained from a blood sample collected from the child during the child's admission in hospital. It is most often the first blood sample collected for several tests. [2]Cases with either O1αβ,2α or O1αβ,2β O-type were compared to controls.

**Supplementary Table 7: Geometric mean concentrations (and 95% confidence intervals) between cord blood from infants with and without invasive *Klebsiella pneumoniae* disease for various anti-K, anti-O and MrkA IgG antibodies.**

| Case definition | IgG | Case [RLU/mL] | Control [RLU/mL] | p-value |
|---|---|---|---|---|
| K2 | K2 | 520 (290-933) n=19 | 678 (575-799) n=280 | 0.372 |
| K25 | K25 | 521 (323-842) n=22 | 1133 (953-1346) n=280 | 0.004 |
| K102 | K102 | 550 (313-968) n=20 | 606 (524-702) n=280 | 0.734 |
| K149 | K149 | 395 (253-616) n=13 | 499 (440-567) n=280 | 0.294 |
| O1αβ,2α | O1αβ,2α | 1745 (393-7743) n=11 | 2318 (1952-2753) n=280 | 0.686 |
| O1αβ,2β | O1αβ,2β | 980 (560-1712) n=20 | 1803 (1529-2126) n=280 | 0.040 |
| O1αβ,2α or O1αβ,2β[1] | O1αβ,2α | 1375 (750-2522) n=31 | 2318 (1952-2753) n=280 | 0.101 |
| O1αβ,2α or O1αβ,2β[1] | O1αβ,2β | 1111 (617-2002) n=31 | 1803 (1529-2126) n=280 | 0.116 |
| O2β | O2β | 365 (251-532) n=20 | 442 (393-497) n=280 | 0.326 |
| O5 | O5 | 784 (510-1206) n=21 | 1004 (875-1152) n=280 | 0.268 |
| All cases | MrkA | 1016 (782-1322) n=99 | 1607 (1376-1876) n=293 | 0.003 |
| K2 | MrkA | 1108 (745-1649) n=19 | 1641 (1402-1922) n=280 | 0.068 |
| K25 | MrkA | 952 (532-1703) n=22 | 1641 (1402-1922) n=280 | 0.074 |
| K102 | MrkA | 1192 (890-1596) n=20 | 1641 (1402-1922) n=280 | 0.056 |
| K149 | MrkA | 1069 (551-2076) n=13 | 1641 (1402-1922) n=280 | 0.199 |
| O1αβ,2α | MrkA | 1043 (519-2094) n=11 | 1641 (1402-1922) n=280 | 0.192 |
| O1αβ,2β | MrkA | 1108 (704-1744) n=20 | 1641 (1402-1922) n=280 | 0.103 |

| | | | | |
|---|---|---|---|---|
| O2β | MrkA | 1172 (874-1570) n=20 | 1641 (1402-1922) n=280 | 0.045 |
| O4 | MrkA | 414 (4-46916) n=2 | 1641 (1402-1922) n=280 | 0.428 |
| O5 | MrkA | 1065 (608-1866) n=21 | 1641 (1402-1922) n=280 | 0.137 |

[1]Cases with either O1αβ,2α or O1αβ,2β O-type were compared to controls.

**Supplementary Figure 1: Correlation between maternal IgG, cord:maternal ratio and infant IgG with birth weight (y-axis is different for each sub-figure to highlight the trends for the different IgG).** Alt text: Graphical representation of estimated maternal, Cord:maternal ratio and cord anti-K and anti-O IgG along with confidence intervals by birth weight in subfigures a to c, respectively.

a)

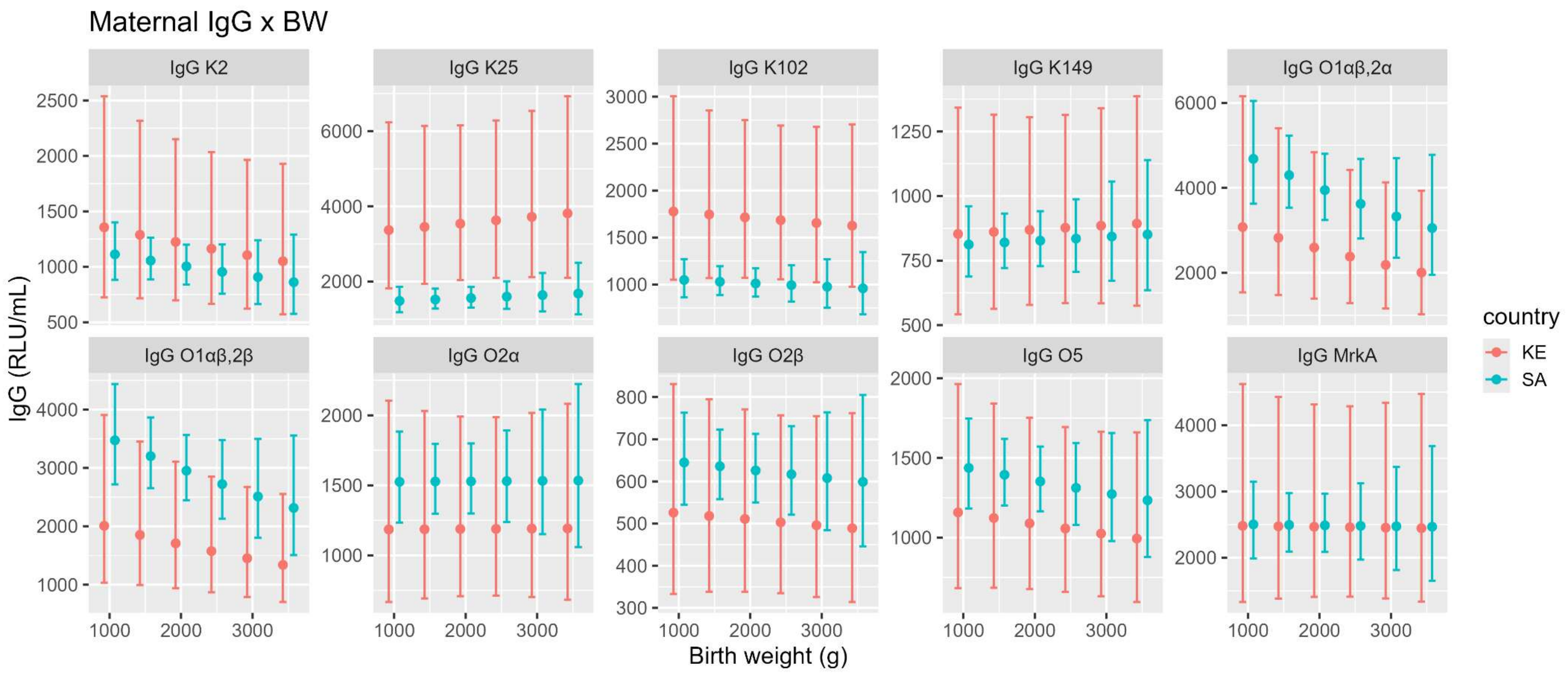

b)

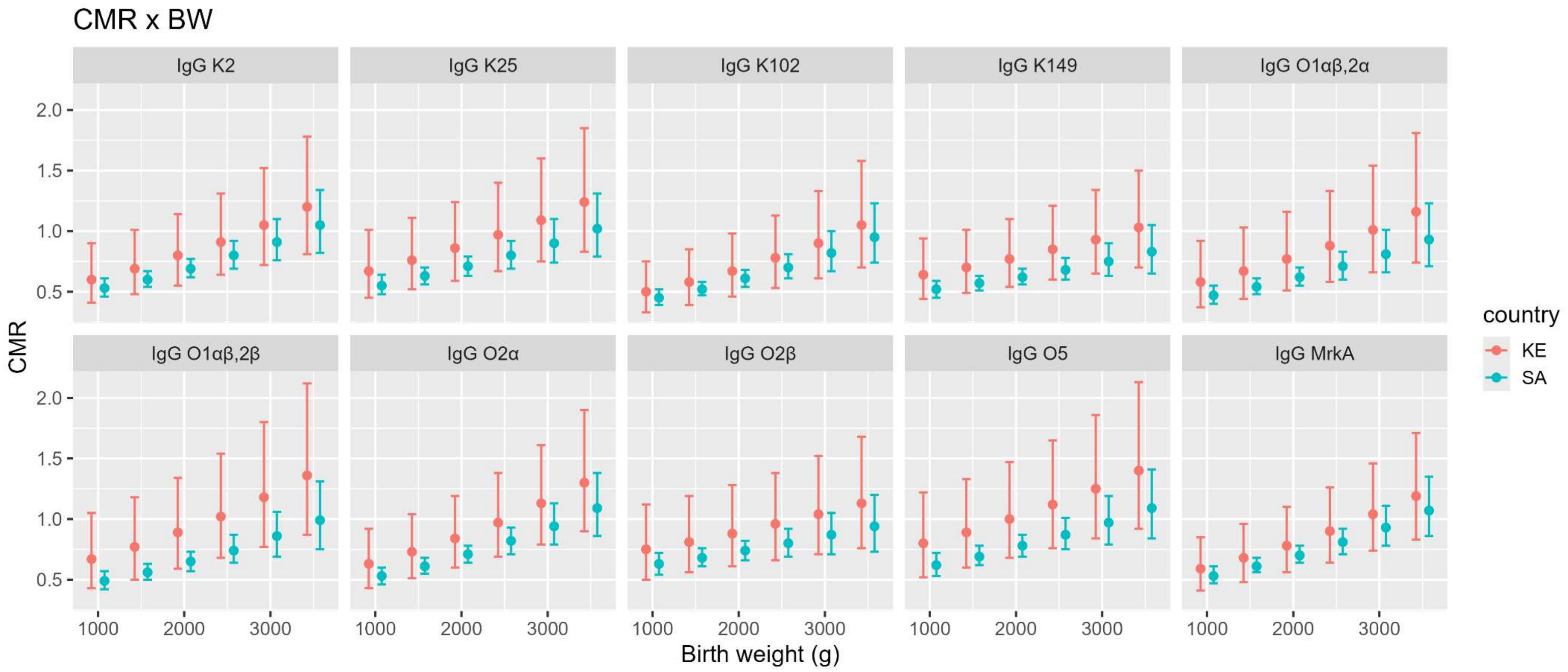
CMR x BW
IgG K2
IgG K25
IgG K102
IgG K149
IgG O1αβ,2α
IgG O1αβ,2β
IgG O2α
IgG O2β
IgG O5
IgG MrkA
CMR
2.0
1.5
1.0
0.5
1000
2000
3000
Birth weight (g)
country
KE
SA

c)

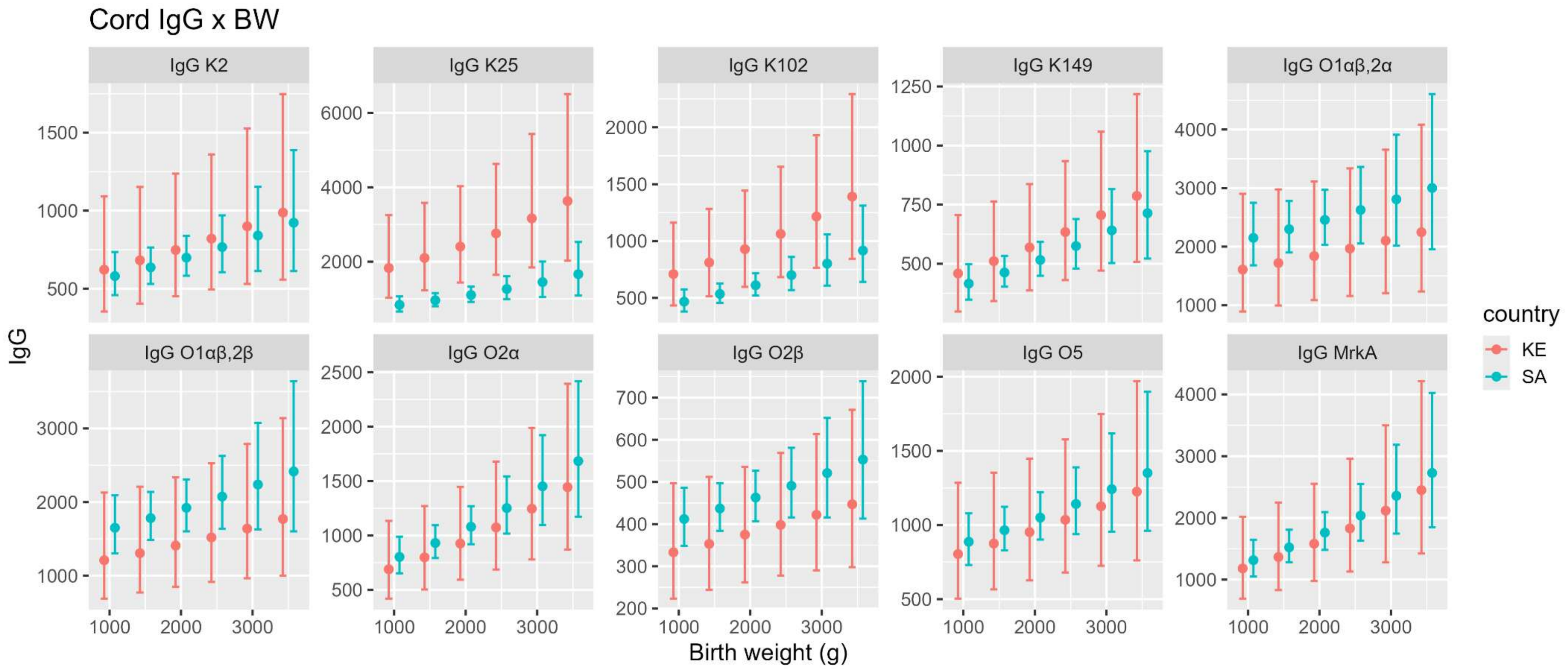

**Supplementary Figure 2: Association between cord-maternal ratio of serum bactericidal assay for K2, K25, K102 and K149 with (a) birth weight and (b) gestational age.** Alt text: Graphical representation of estimated Cord:maternal ratio of SBA along with confidence intervals by birth weight and gestational age in subfigures a and b, respectively.

**a)**

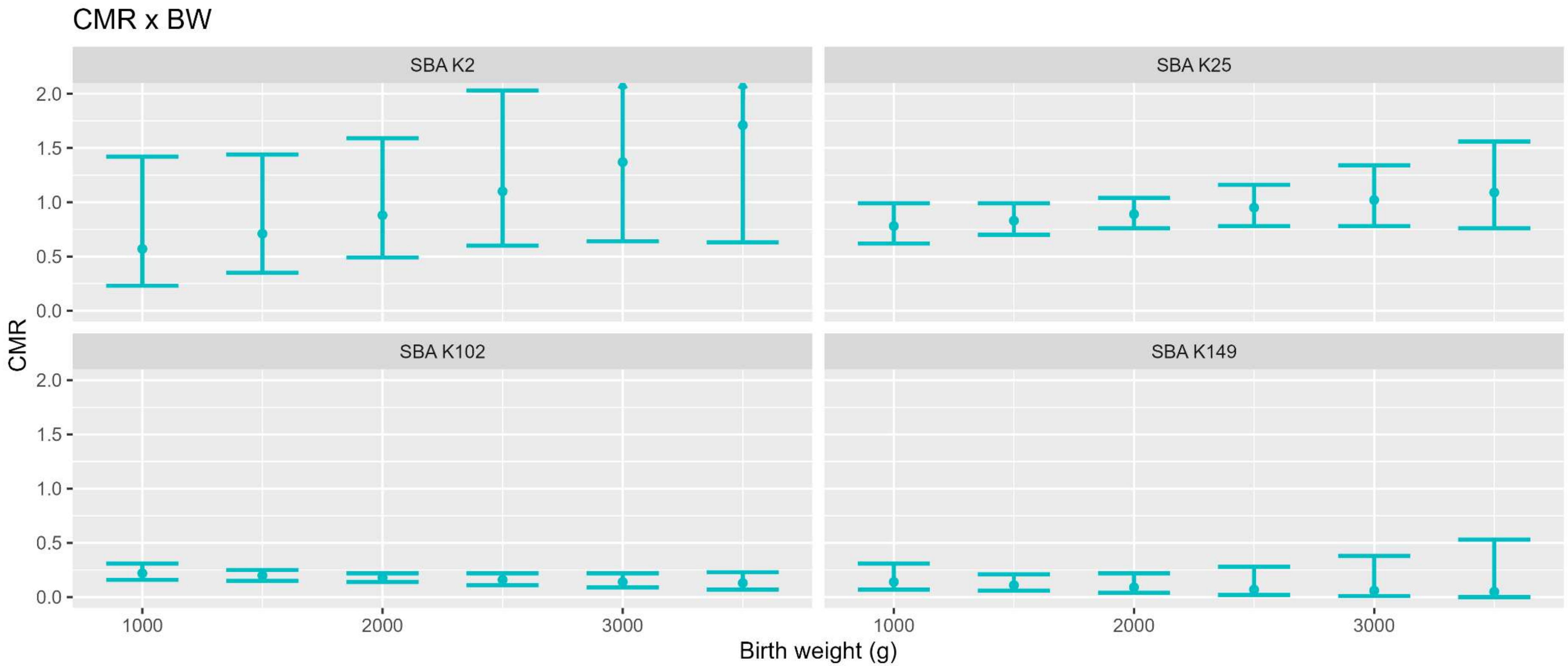
CMR x BW
SBA K2
SBA K25
SBA K102
SBA K149
CMR
2.0
1.5
1.0
0.5
0.0
1000
2000
3000
Birth weight (g)

**b)**

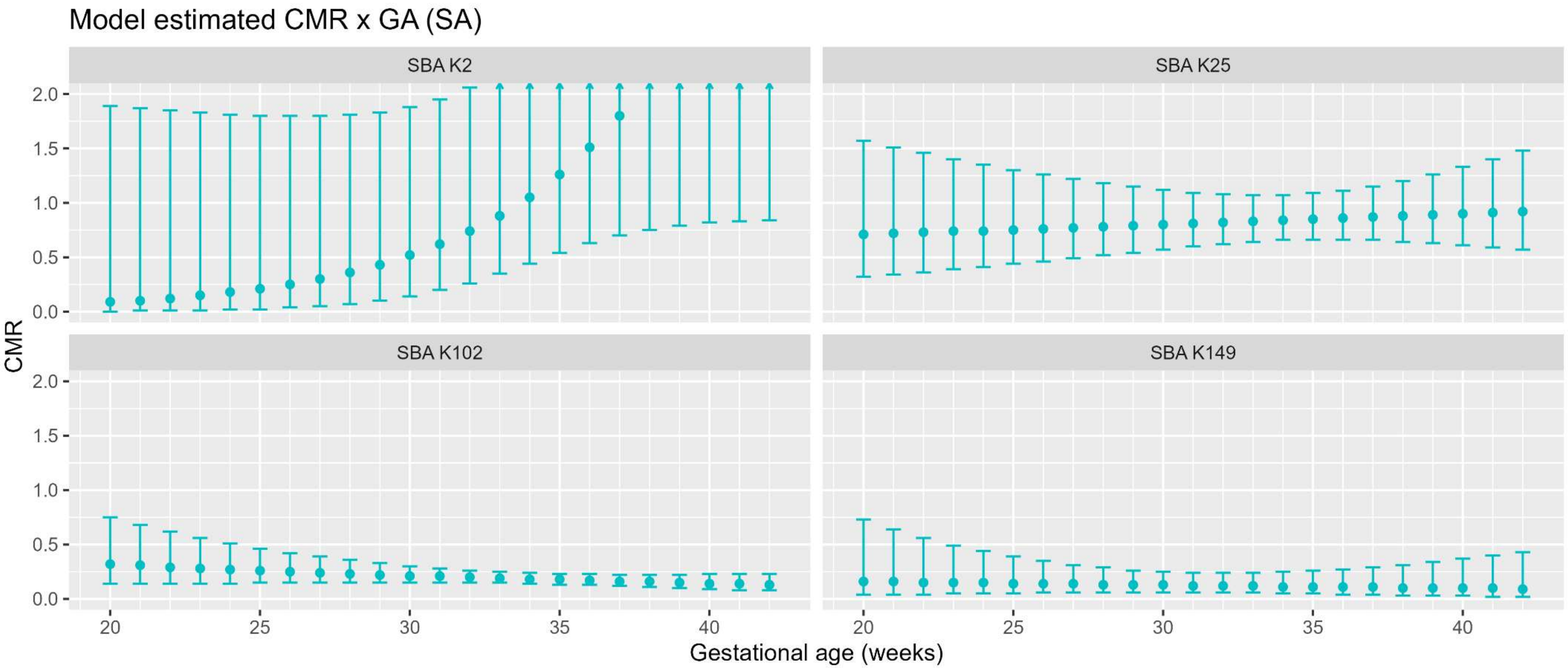
Model estimated CMR x GA (SA)
SBA K2
SBA K25
SBA K102
SBA K149
CMR
0.0
0.5
1.0
1.5
2.0
20
25
30
35
40
Gestational age (weeks)

**Supplementary Figure 3: Reverse cumulative distribution of anti-K (a), anti-O (b) and MrkA (c) IgG in infant cases and controls.** Alt text: Graphical representation of reverse cumulative distribution of anti-K, anti-O and MrkA IgG in cases and controls in subfigures a to c, respectively.

a)

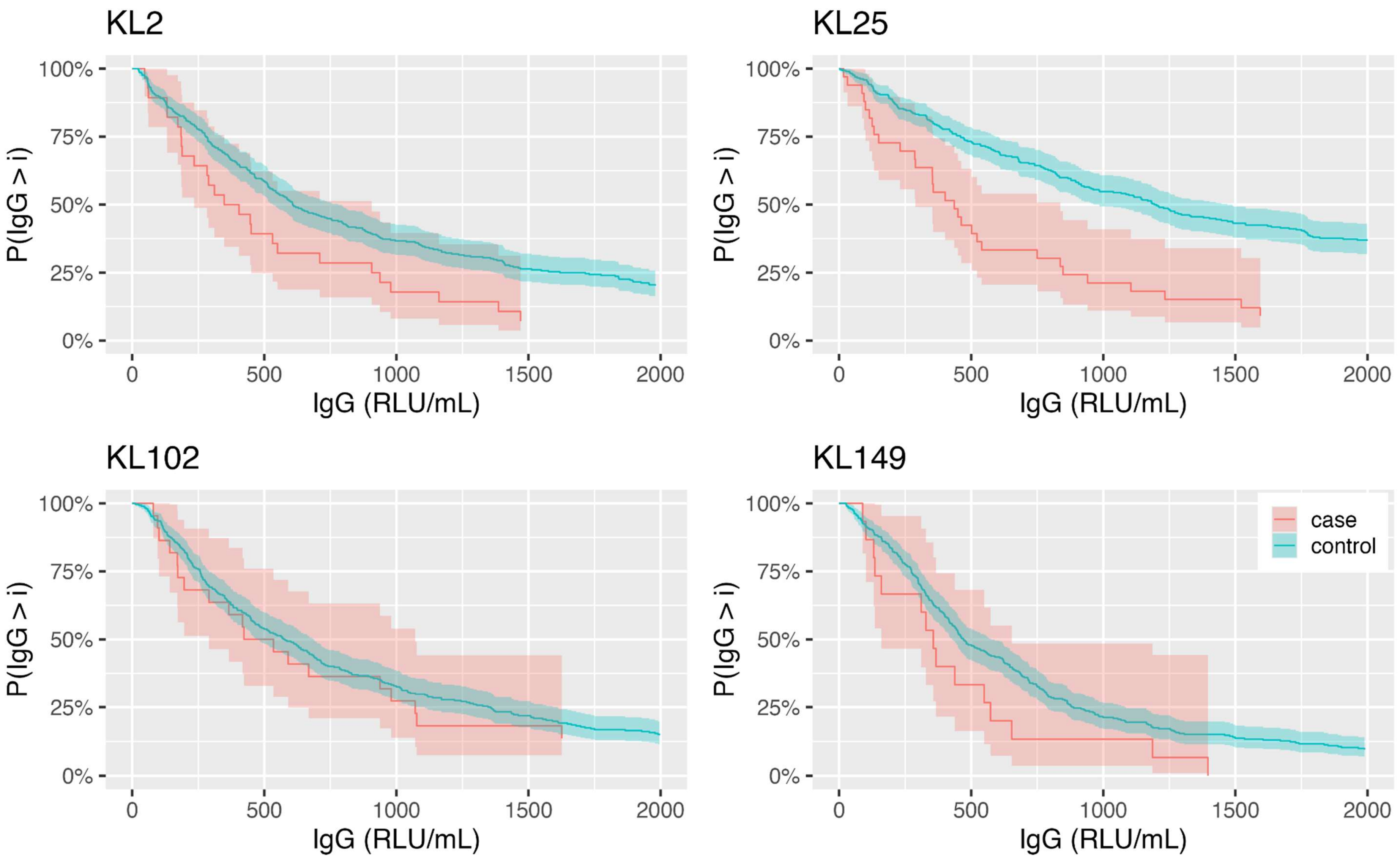
KL2
KL25
KL102
KL149
100%
75%
50%
25%
0%
P(IgG > i)
0
500
1000
1500
2000
IgG (RLU/mL)
case
control

b)

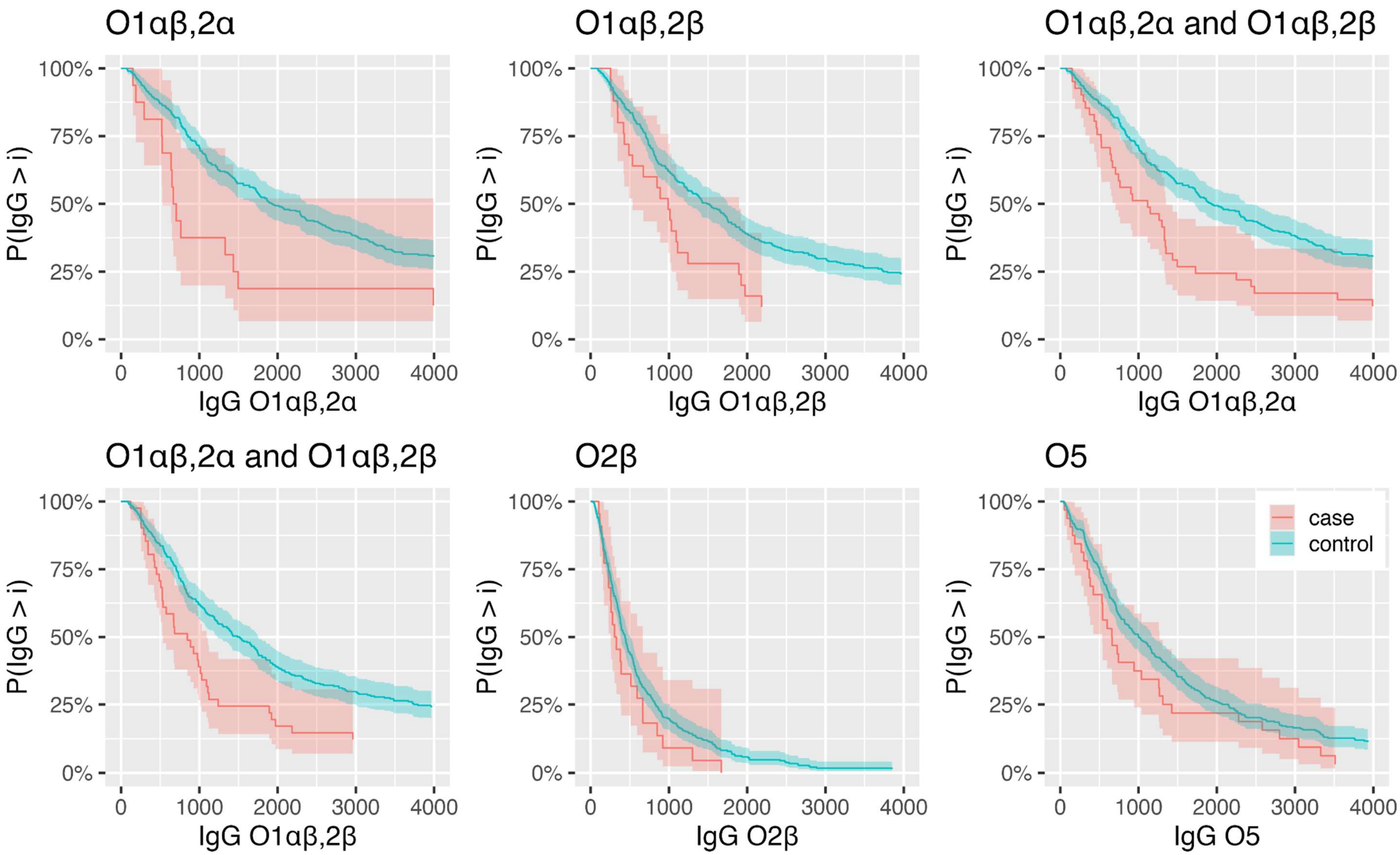

O1αβ,2α
O1αβ,2β
O1αβ,2α and O1αβ,2β
O1αβ,2α and O1αβ,2β
O2β
O5
P(IgG > i)
100%
75%
50%
25%
0%
0
1000
2000
3000
4000
IgG O1αβ,2α
IgG O1αβ,2β
IgG O1αβ,2α
IgG O1αβ,2β
IgG O2β
IgG O5
case
control

c)

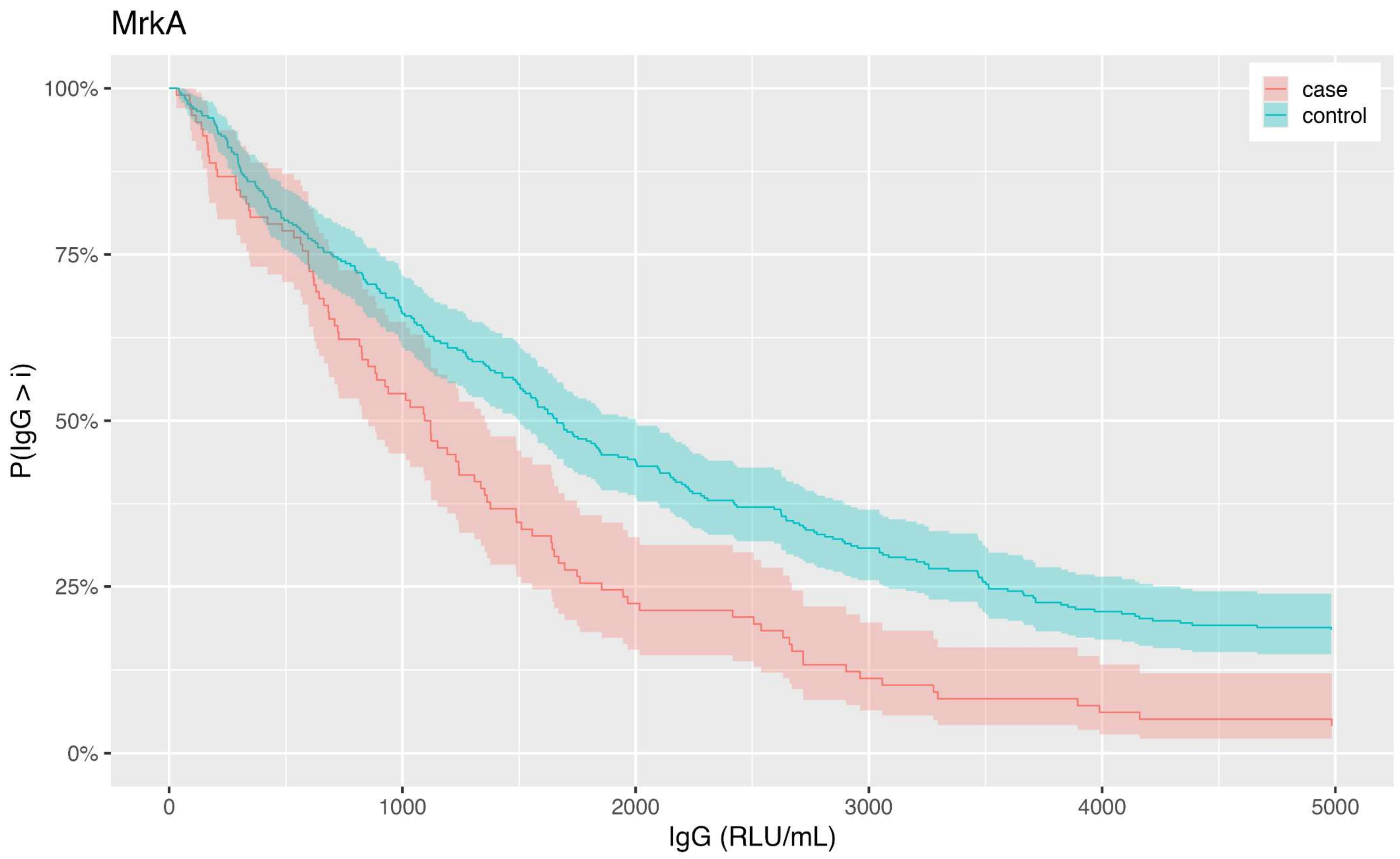
MrkA
100%
75%
50%
25%
0%
P(IgG > i)
0
1000
2000
3000
4000
5000
IgG (RLU/mL)
case
control

**Figure 4: Dot plot with overlayed box-plots (whiskers extend to 1.5 times interquartile-range) of serum bactericidal assay values for K serotypes (a) and O serotypes (b) without values below the lower limit of quantification in cases and controls.** Alt text: Graphical representation of SBA for K and O serotypes in cases and controls using dot plots with overlaying box and whisker-plots in subfigures a and b, respectively.

**a)**

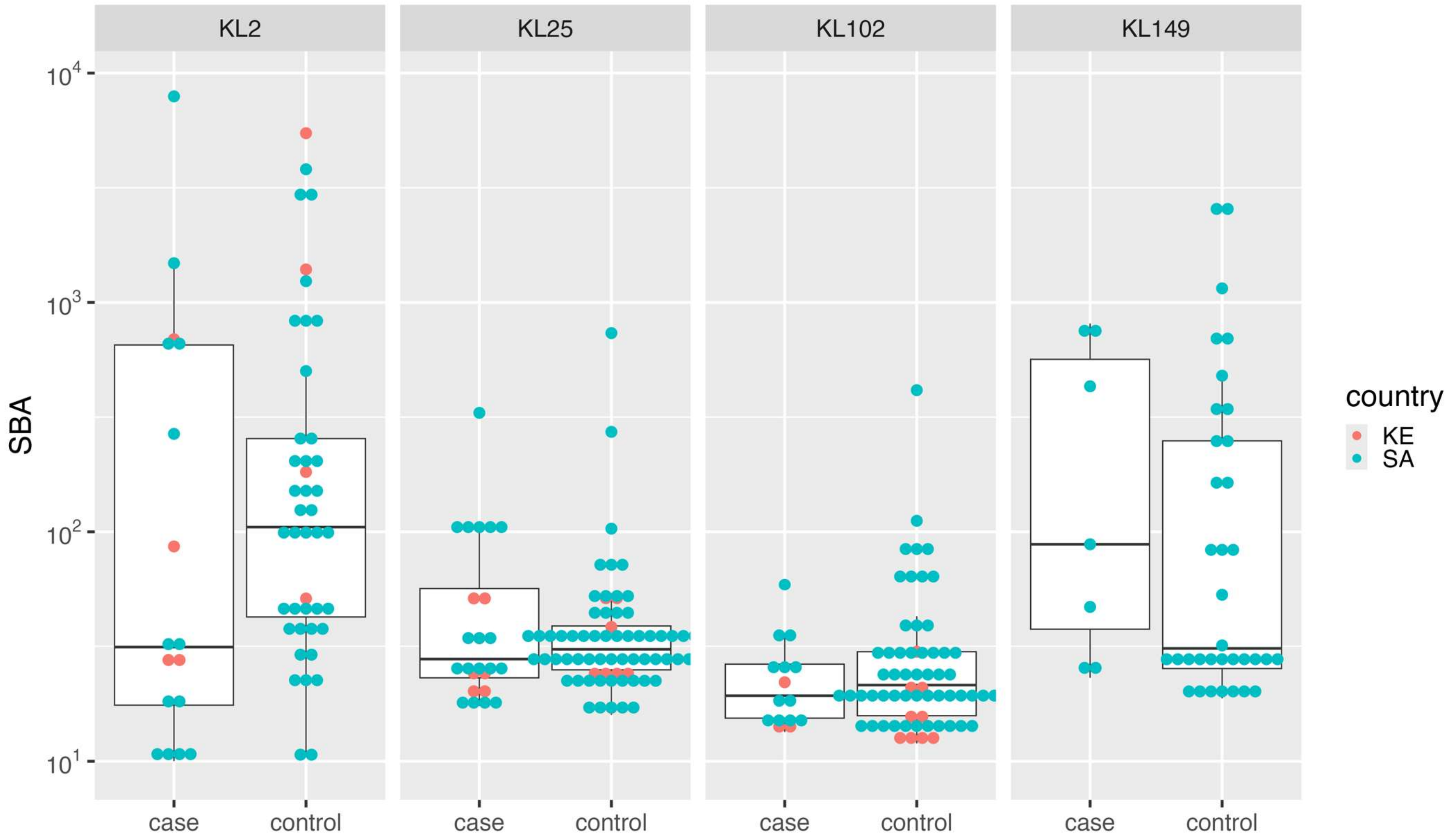
KL2
KL25
KL102
KL149
SBA
$10^4$
$10^3$
$10^2$
$10^1$
case
control
case
control
case
control
case
control
country
KE
SA

**b)**

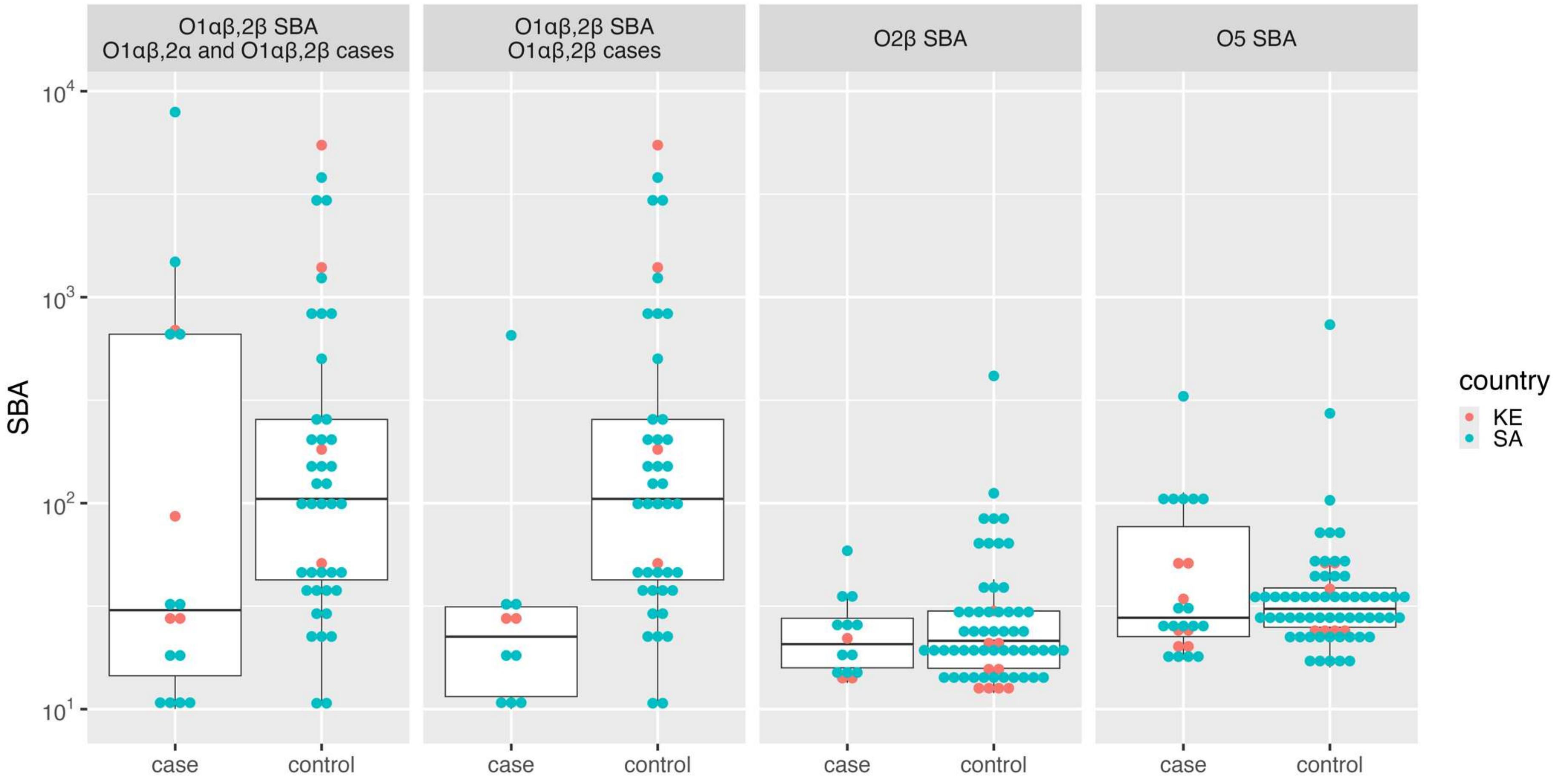

**Supplementary Figure 5: Correlation between serum bactericidal activity and anti-K IgG including (a) and excluding (b) values below lower limit of quantification; and anti-O including (c) and excluding (d) values below lower limit of quantification; and**

**MrkA IgG  including (e) and excluding (f) values below lower limit of quantification.  Correlation estimates are presented within figures.** Points lying on the horizontal line at the minimum log SBA value indicate measurements below the lower limit of quantification for SBA.   Alt text: Graphical representation of correlation between SBA and IgG for anti-K, anti-O and MrkA including and excluding values below the lower limit of quantification in subfigures a to f.

a)

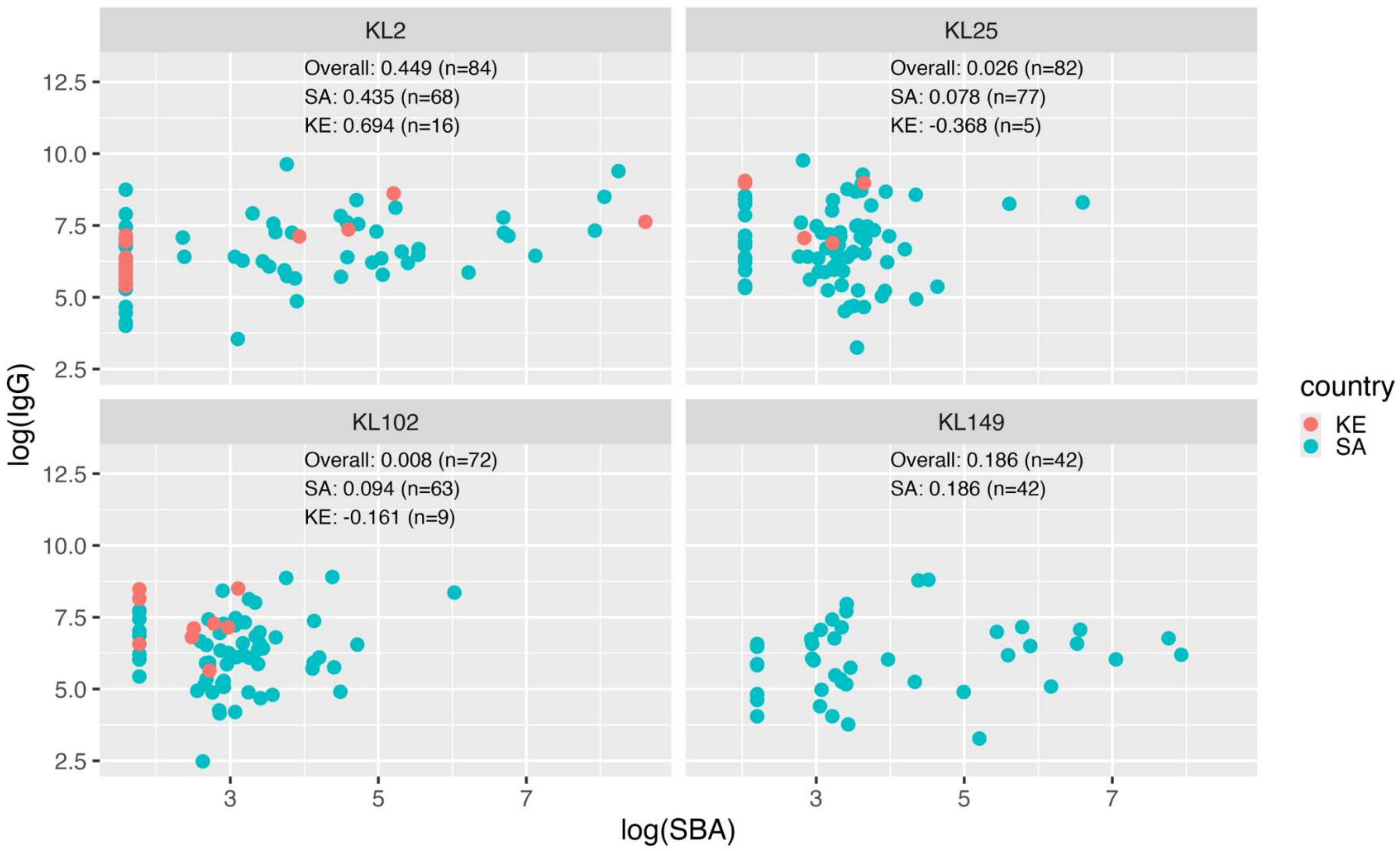

KL2
Overall: 0.449 (n=84)
SA: 0.435 (n=68)
KE: 0.694 (n=16)
KL25
Overall: 0.026 (n=82)
SA: 0.078 (n=77)
KE: -0.368 (n=5)
KL102
Overall: 0.008 (n=72)
SA: 0.094 (n=63)
KE: -0.161 (n=9)
KL149
Overall: 0.186 (n=42)
SA: 0.186 (n=42)
log(IgG)
12.5
10.0
7.5
5.0
2.5
3
5
7
log(SBA)
country
KE
SA

b)

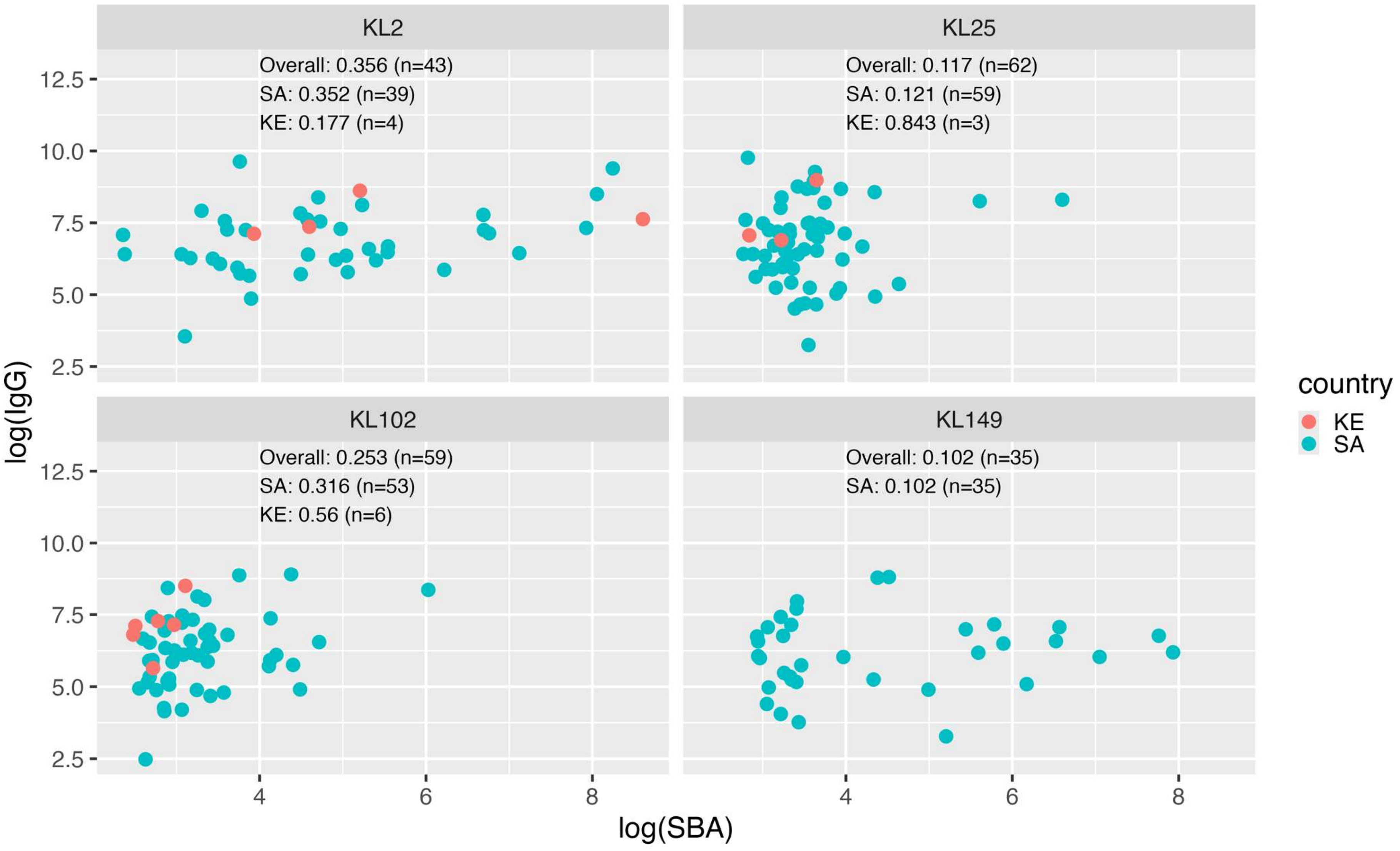
KL2
Overall: 0.356 (n=43)
SA: 0.352 (n=39)
KE: 0.177 (n=4)
KL25
Overall: 0.117 (n=62)
SA: 0.121 (n=59)
KE: 0.843 (n=3)
KL102
Overall: 0.253 (n=59)
SA: 0.316 (n=53)
KE: 0.56 (n=6)
KL149
Overall: 0.102 (n=35)
SA: 0.102 (n=35)
log(IgG)
12.5
10.0
7.5
5.0
2.5
4
6
8
log(SBA)
country
KE
SA

c)

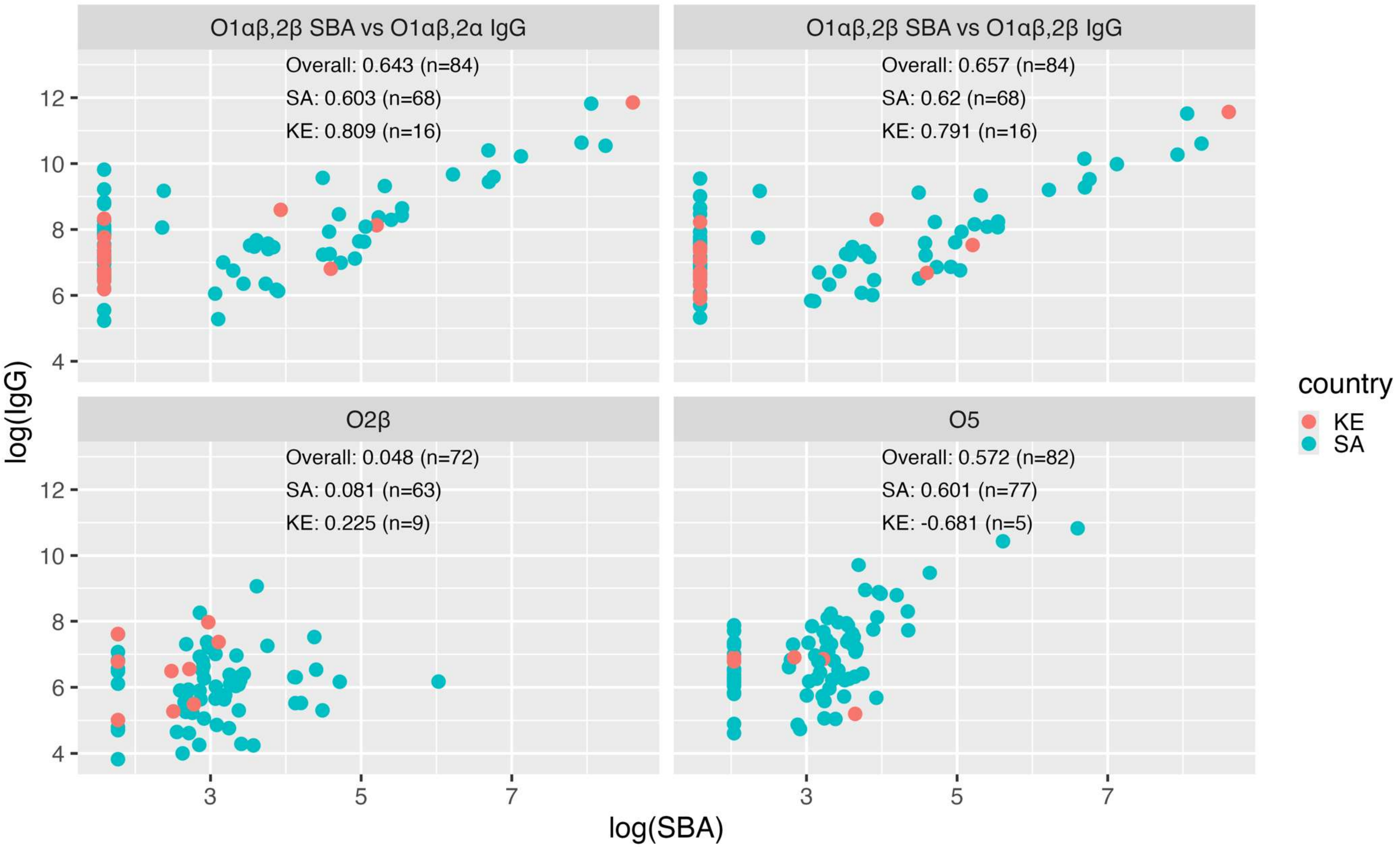

O1αβ,2β SBA vs O1αβ,2α IgG
Overall: 0.643 (n=84)
SA: 0.603 (n=68)
KE: 0.809 (n=16)
O1αβ,2β SBA vs O1αβ,2β IgG
Overall: 0.657 (n=84)
SA: 0.62 (n=68)
KE: 0.791 (n=16)
O2β
Overall: 0.048 (n=72)
SA: 0.081 (n=63)
KE: 0.225 (n=9)
O5
Overall: 0.572 (n=82)
SA: 0.601 (n=77)
KE: -0.681 (n=5)
log(IgG)
log(SBA)
country
KE
SA

d)

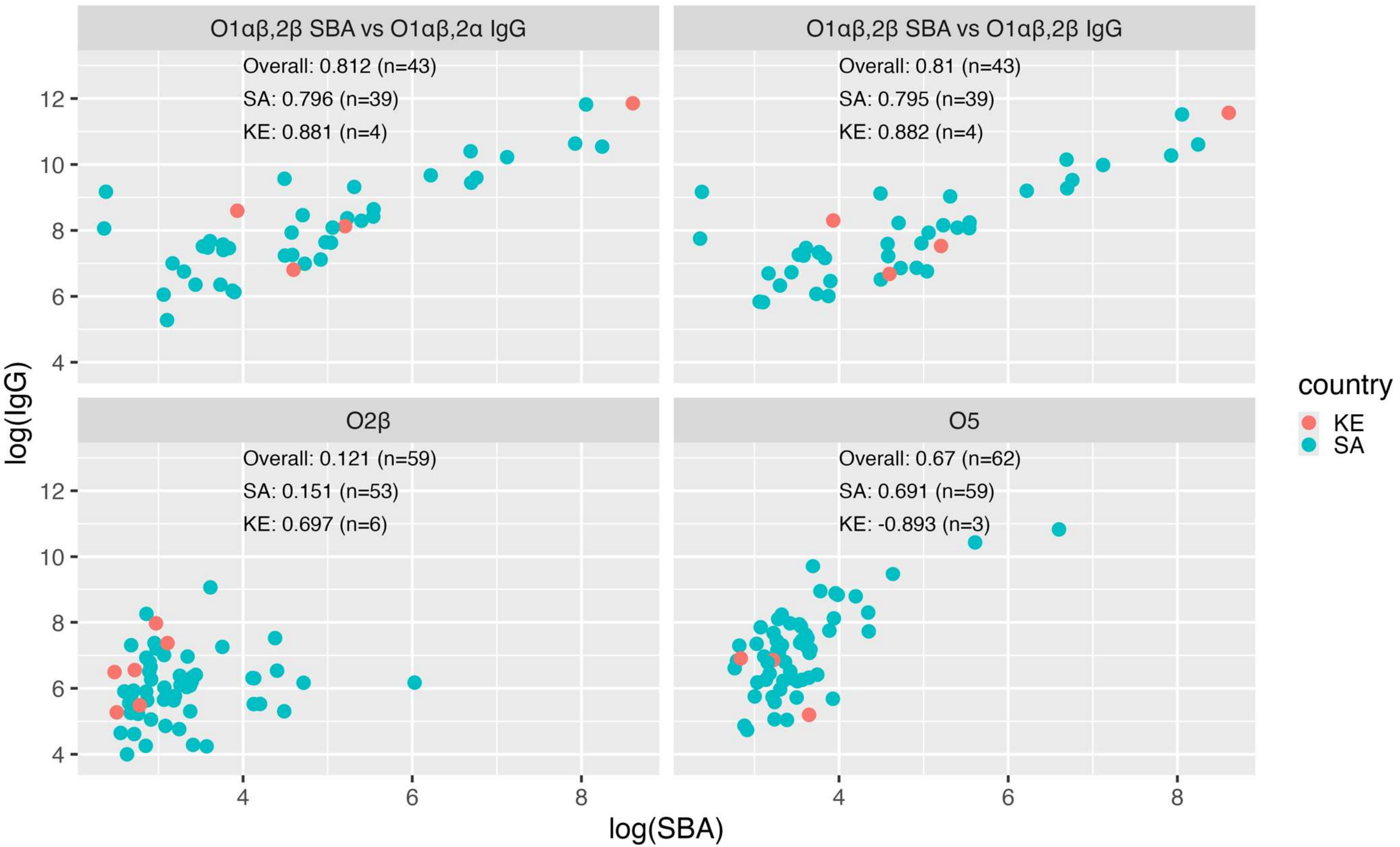
O1αβ,2β SBA vs O1αβ,2α IgG
Overall: 0.812 (n=43)
SA: 0.796 (n=39)
KE: 0.881 (n=4)
O1αβ,2β SBA vs O1αβ,2β IgG
Overall: 0.81 (n=43)
SA: 0.795 (n=39)
KE: 0.882 (n=4)
O2β
Overall: 0.121 (n=59)
SA: 0.151 (n=53)
KE: 0.697 (n=6)
O5
Overall: 0.67 (n=62)
SA: 0.691 (n=59)
KE: -0.893 (n=3)
log(IgG)
log(SBA)
country
KE
SA

**e)**

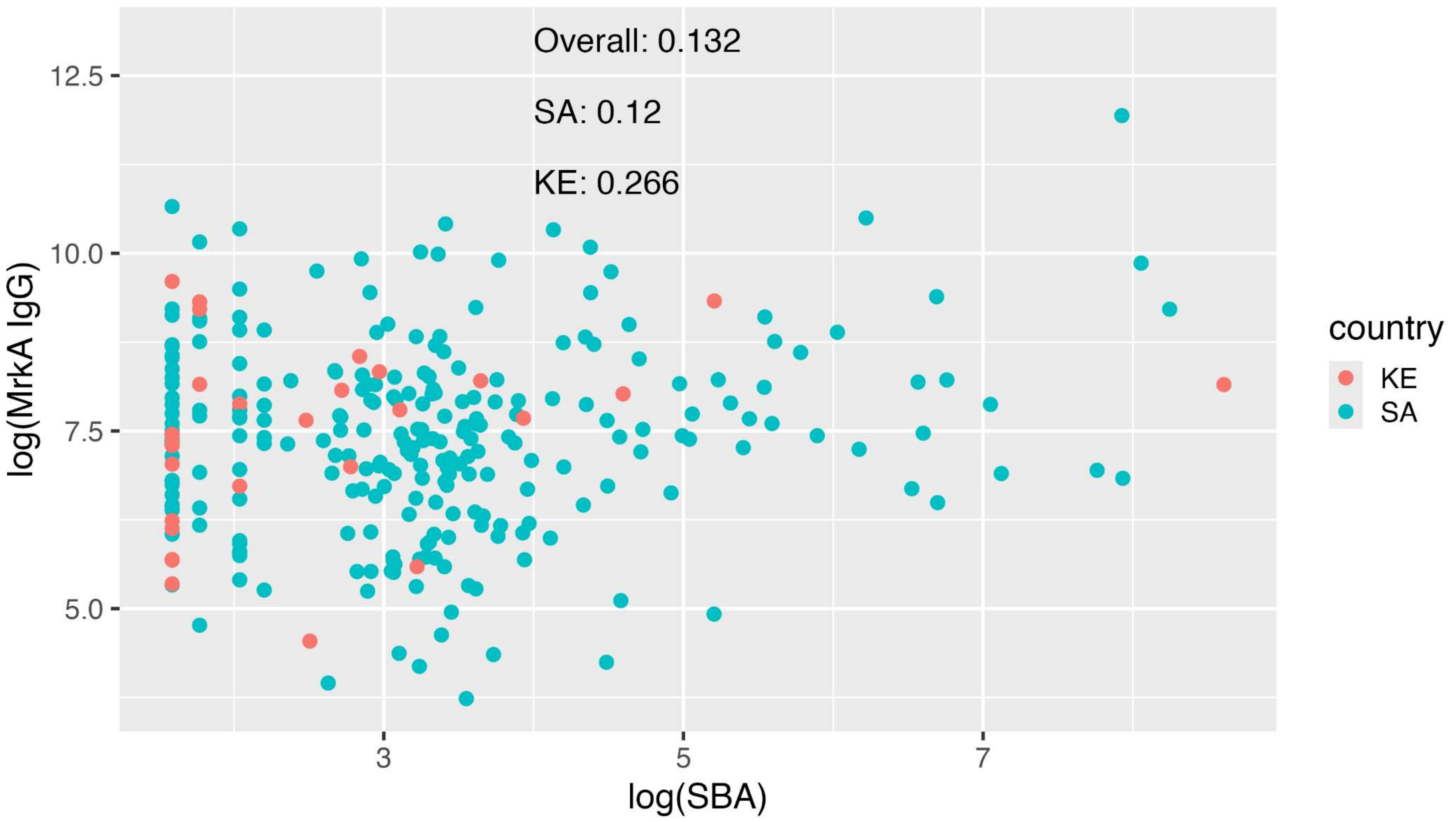
Overall: 0.132
SA: 0.12
KE: 0.266
12.5
10.0
7.5
5.0
log(MrkA IgG)
3
5
7
log(SBA)
country
KE
SA

f)

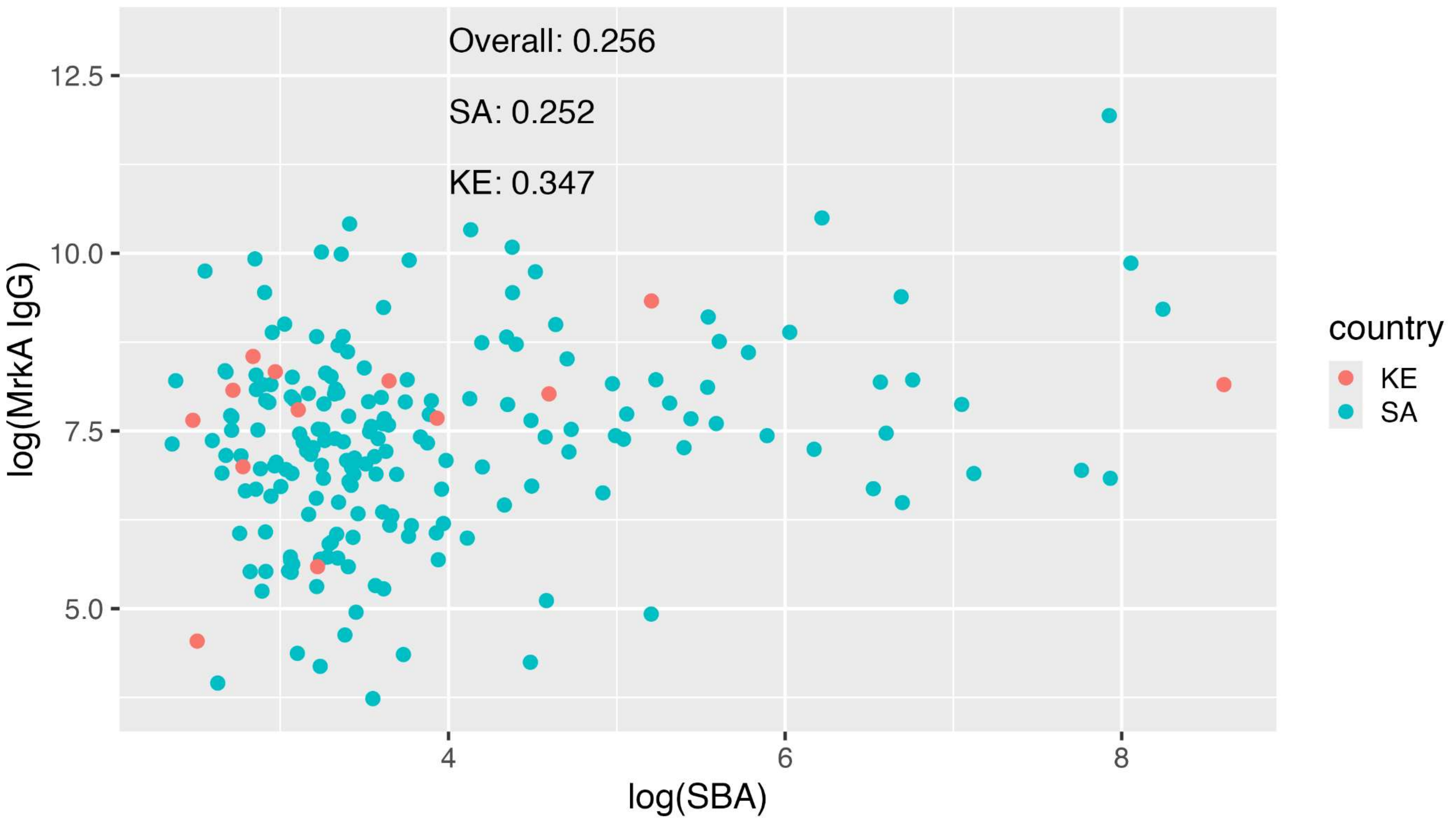

Overall: 0.256
SA: 0.252
KE: 0.347
log(MrkA IgG)
12.5
10.0
7.5
5.0
4
6
8
log(SBA)
country
KE
SA
